# How do Urban Factors Control the Severity of COVID-19?

**DOI:** 10.1101/2022.06.17.22276576

**Authors:** Jacob Roxon, Marie-Sophie Dumont, Eric Vilain, Roland J.M. Pellenq

**Affiliations:** EpiDaPo Lab - CNRS / George Washington University Children’s National Medical Center, Children’s Research Institute, 111 Michigan Ave. NW, Washington, DC 20010, USA

## Abstract

Human health in urban environment has emerged as a primary focus of sustainable development during the time of global pandemic caused by a severe acute respiratory syndrome due to the SARS-CoV-2 coronavirus, COVID-19. It has reshaped the world with the way our communities interact, people work, commute, and spend their leisure time. While different mitigation solutions for controlling COVID-19 virus transmission have already been established, global models that would explain and predict the impact of urban environments on the case fatality ratio *CFR* of COVID-19 (defined as the number of deaths divided by the number of cases over a time window) are missing. Here, with readily available data from public sources, we study the *CFR* of the coronavirus for 118 locations (city zip-codes, city boroughs, and cities) worldwide to identify the links between the *CFR* and outdoor, indoor and personal urban factors. We show that a probabilistic model, optimized on the sample of 20 districts from 4 major US cities, provides an accurate predictive tool for the *CFR* of COVID-19 regardless of the geographical location. When adjusted for the population, our model can be used to evaluate risk and severity of the disease at multi-geospatial scales worldwide ranging from zip-codes and neighborhoods to cities and countries for different waves of the pandemic. Our results suggest that although disease screening and vaccination policies to containment and lockdowns remain critical in controlling the spread of airborne diseases, urban factors such as population density, humidity, or order of buildings, should all be taken into consideration when identifying resources and planning targeted responses to mitigate the impact and severity of the viruses transmitted through air. We advocate the study of urban factors as a path towards facilitating timely deployment of targeted countermeasures and confinement strategies where sharing of personal information and availability of tests may be restricted or limited.

## Introduction

Towards the end of January 2020, the World Health Organization (WHO) declared a Public Health Emergency of International Concern, which within less than 6 weeks, on March 11th 2020 was described as a global pandemic of coronavirus (COVID-19) disease. The most severe impact of the COVID-19 has been observed in urban areas (1, 2), which in most countries are home to over 70% of population. And while the implementation of pandemic response measures, such as vaccinations or social distancing and face covering restrictions, have provided a path towards controlling the epidemic (3, 4), they may not be sufficient to end the disease, which is likely to become endemic (5) resulting in infections and mortalities, severity of which, seen through the prism of the Case Fatality Ratio, *CFR*, (i.e. the ratio of the number of deaths to the number of confirmed cases) changes from country to country (Fig. 1).

**Figure 1.**
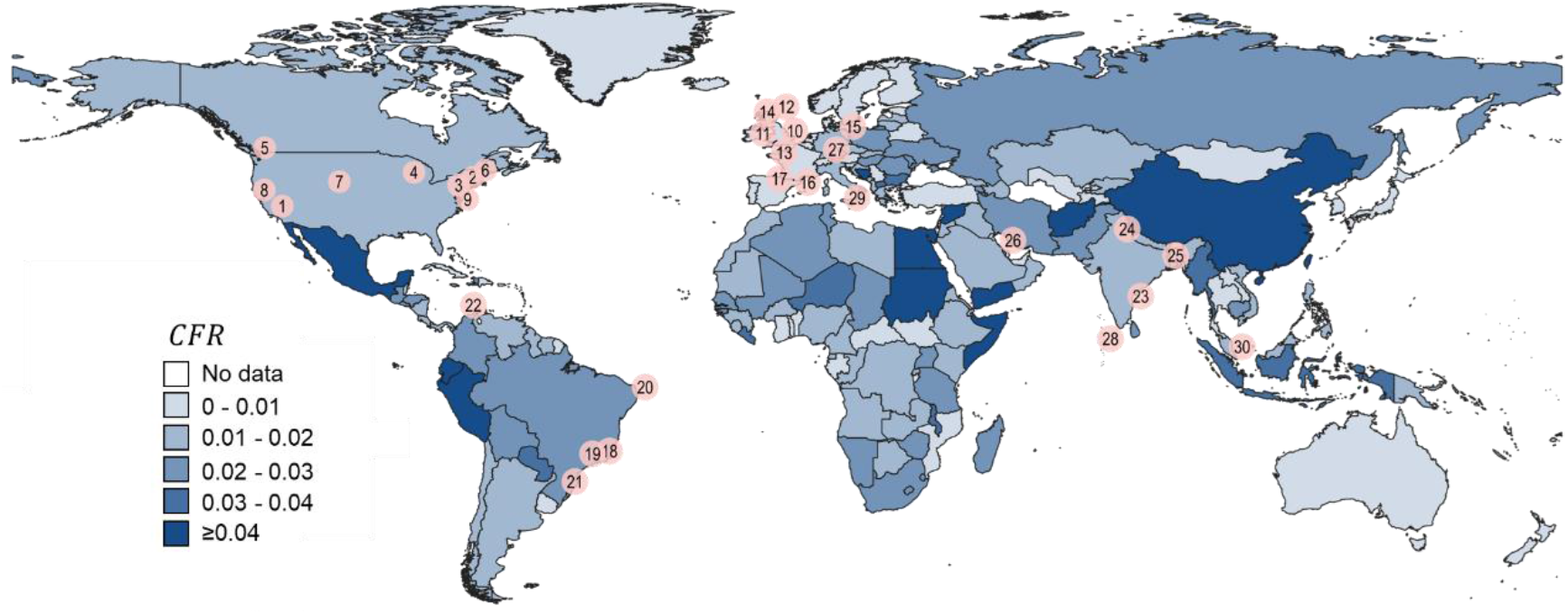
The severity of the COVID-19 throughout the world showing cumulative COVID-19 *CFR*(Table S1) at the country scale between 01/2020 and 01/2022 with a geo-location of 30 cities (Table S2) used in this study.

SARS-CoV-2 virus stability is affected by physical, chemical, and biological factors (6). On inanimate surfaces, the most important are surface type, relative humidity (7), temperature (8), moisture content of the suspending medium (mucus). Although among humans, there are different possible transmission routes of COVID-19, the focal point of scientists has been airborne propagation of the SARS-CoV-2 (9), which involves projection of virions in aerosolized droplets of mucus from an infected individual, inhaled by sane individual. Due to air drag and gravity, virus-laden droplets with diameter, *d* ≥ 5 µ*m* emitted in a cough or sneeze, have a generally low projection distance of less than few meters (10, 11). Smaller virus-laden particles (*d* < 5 µ*m*), can remain in air for hours and consequently be dispersed through airborne route both indoors and outdoors. In addition, the propagation of COVID-19 has been linked to particulate matter (PM) pollution levels showing a positive correlation between long-term exposure to high concentration of PM particles and the number of COVID-19 deaths (12). At a nanoscale, the viral load and virulence through the SARS-CoV-2 spike proteins and ACE2 human cells receptors fusion mechanisms, correlate directly with infectivity, disease phenotype, morbidity, and mortality, which should be accounted for correlating conventional aerosol metrics, such as PM2.5 or PM10 concentrations to COVID-19 cases, or deaths, and provide a direct explanation of the variability of the COVID-19 severity through the Case Fatality Ratio (13). *CFR* has also been linked to the viremia (14, 15), a measure of the quantity of virions emitted by infected patients in the form of aerosolized droplets that can circulate both indoors and outdoors. Although, *CFR* is considered a crucial quantity to predict emergency and health care infrastructure deployment to manage the spread of COVID-19, understanding of mechanisms that in an urban setting contribute to the COVID-19 severity and the airborne propagation of SARS-CoV-2 remains limited, subsequently preventing accurate determination of COVID-19 *CFR* at the scale of neighborhoods, cities or even countries (16–22). Here, we propose a probabilistic model to predict *CFR* based on 3 distinctive Urban Factors (*U*_*i*_): Personal (*P*_*i*_), Indoor (*I*_*i*_), and Outdoor (*O*_*i*_). We find that while the approach works best for city-wide predications (*N* = 30 cities worldwide), we can also successfully utilize it at other scales, ranging from zip codes or city districts (Fig. S1) to county or even country levels.

## Results

The main component of the *CFR* model relies on the data from the first year of the pandemic for 30 cities worldwide (Fig. 1, Table S1) at various geospatial scales. Since at the very beginning of the pandemic there were major challenges in reporting values of infected people, *C*19_*c*_, we chose a minimal time window of six months, which is sufficient to study the first wave of the COVID-19 virus and investigate *CFR* correlations between *C*19_*c*_ and the number of deaths, *C*19_*d*_ in the form of:

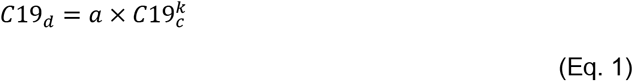

where, *a* is an algebraic proportionality constant and *k* is an exponent factor, which for COVID-19 has been ≅ 1 (Fig. S2). We began evaluating *a* = *CFR*= *C*19_*d*_/*C*19_*c*_ at a local scale for city wards and districts (Fig. S1, Table S2-S3) in 4 US cities: New-York NY (Figs. 2a-b), Los Angeles CA, Chicago IL and Seattle WA, during the first COVID-19 wave (*λ*_1_ factor) from March 2020. Although, similar data for the wards of Washington D.C. is available and has been used throughout this study, we chose not to use it in the model optimization process due to low (<100,000) population of D.C. wards but rather keep this information for validation purpose at the wards scale. While for most regions available data (Table S2) extends beyond the first 6-12 months of the pandemic and captures multiple waves, herein defined *λ*_1_ captures the earliest and most severe time frame of the pandemic. Upon investigating time series of *CFR*s, we find that during *λ*_1_ they resemble sigmoid functions. To eliminate time component, we use average *CFR*(*λ*_1_) obtained directly from upper plateau range of *CFR* distribution (Fig. 2a). Although, one can use the steepest slope (Fig. 2b) or the maximum value (Fig. 2a) of *CFR* distribution to quantify its severity cities districts and cities (*N* = 50, Figs. 2c-d), *CFR*(*λ*_1_) provides the most stable averaging approach that works with inconstancies and fluctuations in raw COVID-19 data. With such defined *CFR*(*λ*_1_) we proceeded to define Personal (*P*_*i*_), Outdoor (*O*_*i*_) and Indoor (*I*_*i*_) Urban Factors, *U*_*i*_s (Fig. 3a), where *i* represents a geographical location, which to formulate the model is limited to wards or districts of cities.

**Figure 2.**
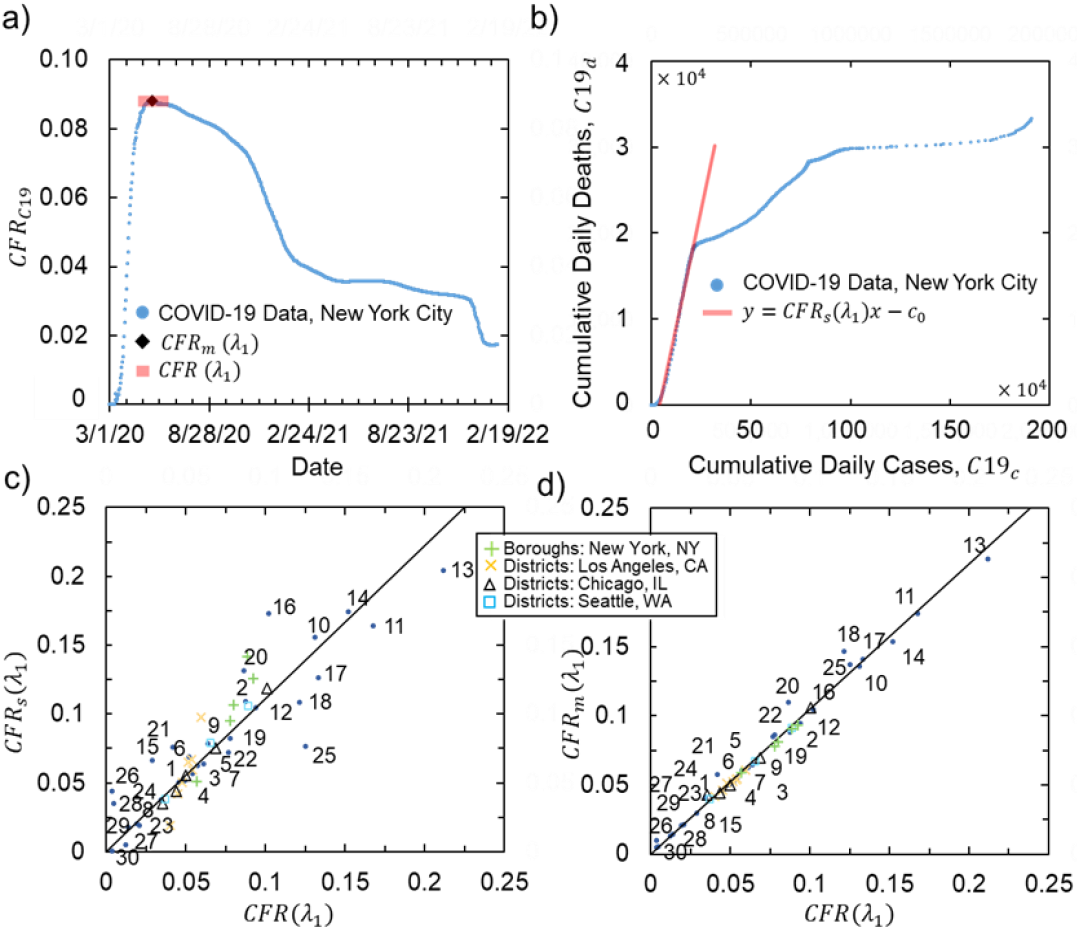
COVID-19 *CFR* methodology showing (a) *CFR* time series with examples for calculating *CFR*(*λ*_1_) and *CFR*_*m*_(*λ*_1_) and (b) *CFR*_*s*_(*λ*_1_) using Eq.1 for the first wave of the pandemic in New York City, NY. Linear correlation between (c) *CFR*(*λ*_1_) vs. *CFR*_*s*_(*λ*_1_) and (d) *CFR*(*λ*_1_) vs. *CFR*_*m*_(*λ*_1_) for city districts (*N* = 20, Table S4) and cities (*N* = 30, Table S5).

**Figure. 3.**
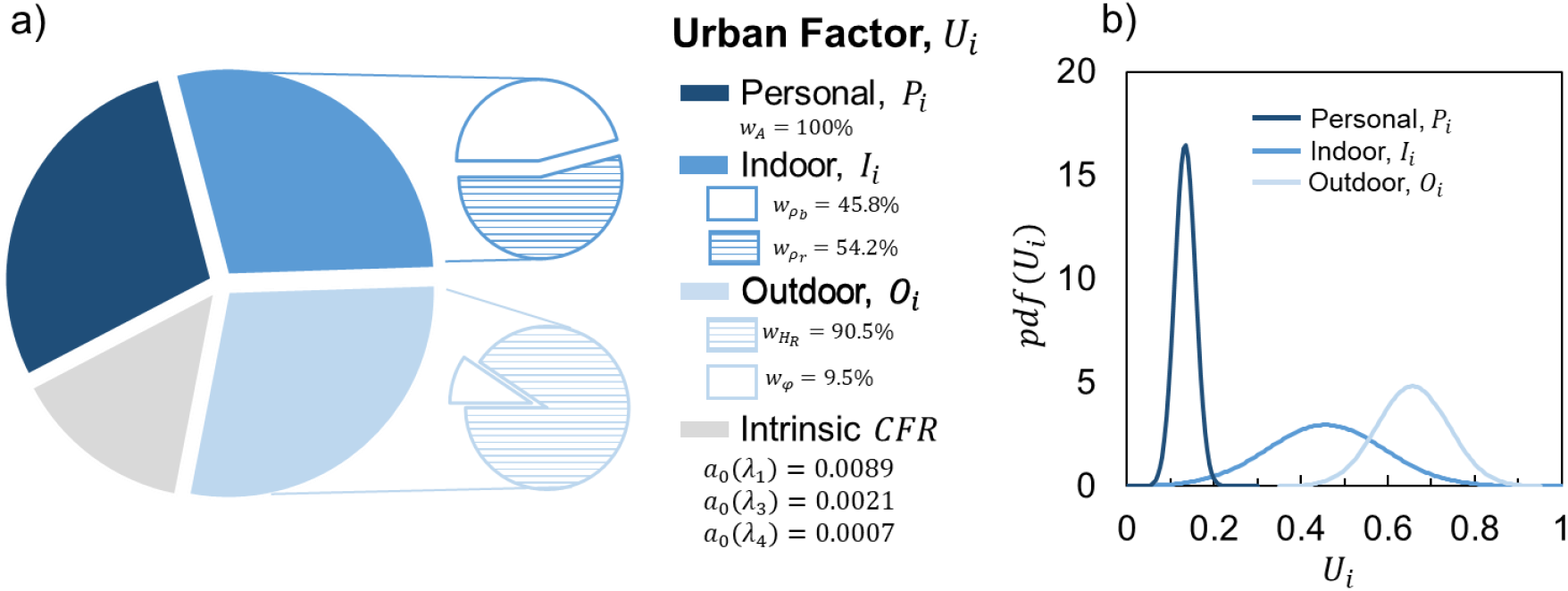
(a) Urban Factors with intrinsic *CFR* values for different waves of the pandemic and model weight parameters optimized for the city districts (SI, Table S4) with (b) their probability distributions.

To identify differences in the COVID-19 spread in each location as captured with *CFR*, we adopted city texture methodology (see Materials and Methods, SI), among other variable used in the determination of Urban Factors (Tables. S4-S8). However, for the calibration of weight parameters in equations 3-5 only city district (*N* = 20) from Table S3 have been used to allow a sufficient sample of uncorrelated data from various geographical regions to be used for validation of the model (*N* = 118 for cities, zip codes and boroughs, including 9 outliers).

To correct measured *CFR* values, the dimensionless weight factors derived from *CFR*(*λ*_1_) for 20 city districts from 4 major US cities were adjusted using a theoretical model *a*_*i*_, defined as the *CFR*(*λ*_1_) for region *i* during time window *λ*_1_ (first pandemic: March-August 2020, Fig. 4a):

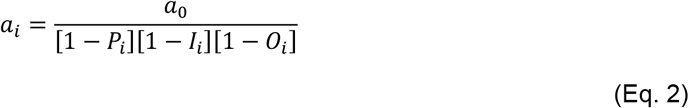

where, *a*_0_ is a constant indicative of intrinsic *CFR*(*λ*_1_) (corrected for the urban factors under the form of a product of 3 independent probabilities) during the initial, first wave of the pandemic with no, or minimal, city-wide social distancing and vaccine protocols. Thus, *a*_0_ becomes *CFR*(*λ*_1_) that corresponds to natural human response to COVID-19 virus corrected for all type of Urban Factors, a correction derived using independent and uncorrelated *U*_*i*_s (Fig. 3b): Personal, Indoor and Outdoor Factors, where each [1 − *U*_*i*_] component reflects the correction of measured *CFR*(*λ*_1_).

**Fig. 4.**
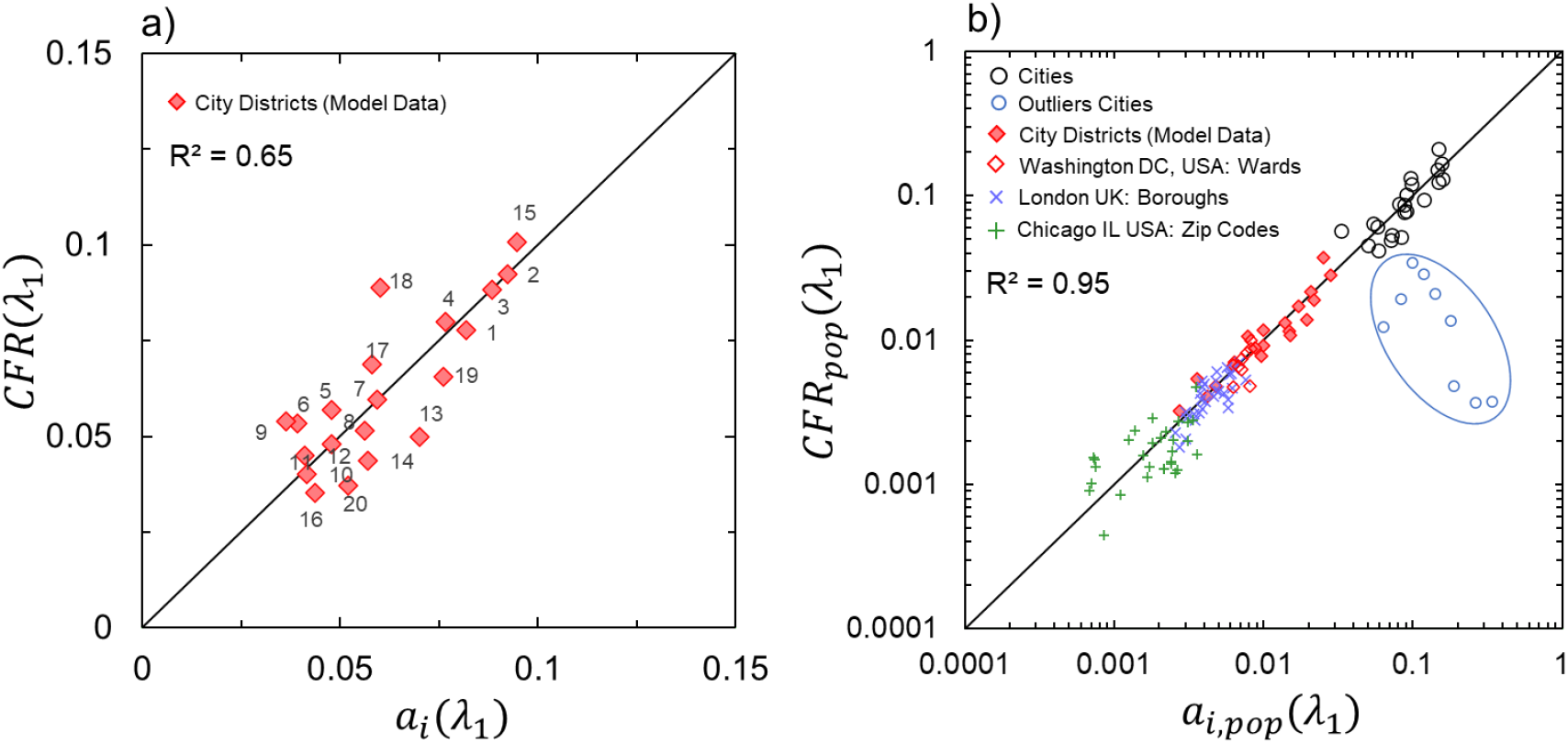
Urban Factors model predictions showing (a) comparison between measured and predicted *CFR*(*λ*_1_) from Eq.2 for 20 city districts for 4 major US cities (Fig. S1, Table S4) between 03/2020-08/2020 used for optimizing urban factor weight parameters and intrinsic *CFR*. Linear fitting with slope coefficient of unity provides *R*^2^= 0.65 and *RMSE* = 0.012. (b) Comparison between measured and predicted population adjusted *CFR*_*pop*_(*λ*_*i*_) for 118 location worldwide (Tables S4-S9) during different waves of the pandemic (excluded 9 outliers for cities in Table S5 for the correlation calculations). Linear fitting with slope coefficient of unity provides *R*^2^= 0.*95* and *RMSE* = 0.31 in the loglog scale. Predicted *CFR* and *CFR*_*pop*_ values used the same urban factor weight parameters in both figures.

From the distribution of factors, it can be concluded that age index, captured by *P*_*i*_, has much lower impact on *CFR*(*λ*_1_) than *O*_*i*_ or *I*_*i*_, where their mean values closer to 1 have greater influence over *CFR*(*λ*_1_). With a very low absolute residual sum between predicted and measured *CFR*, which is < 15% of 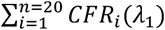 (SI) built upon 20 city districts, the *CFR*(*λ*_1_) model results in *a*_0_ = 0.0089, which at the global scale is lower than previously measured intrinsic *a*_0_(*λ*_1_)= 0.014 (22) for single geographical region. While real-time predictability of *a*_0_ can be improved by considering a day-to-day and local variability of relative humidity, here it is assumed to be an external constraint affecting the whole urban area using annual average values. To adequately capture risk and severity of *CFR*(*λ*_1_) at the local scale of cities, for zip-code, wards, districts and boroughs values we multiple measured and predicted case fatality ratio values by the fraction of the population of a specific city (Fig. 4b). For cities taken as a whole, this ratio remains unity.

## Discussion

The ability of our model to extend beyond city districts scale was then demonstrated at the global city scale with no adjustment of urban factor weight and *a*_0_ parameters for 30 cities and 88 smaller scale locations (i.e. zip-codes, wards) worldwide. Regardless of the scale we find that very accurate population adjusted *CFR*_*pop*_(*λ*_1_) predictions (*R*^2^= 0.95) can be made for any place in the world, provided that no extreme countermeasures were adopted at the beginning of the pandemic. Furthermore, the Urban Factors model from Eq.2 can be applied to make prediction over different time scales of the pandemic. With additional cumulative *C*19_*C*_ and *C*19_*d*_ data between the start of the pandemic and third wave (March 2020 - June 2021) and forth wave (March 2020 - January2022) respectively, and keeping all *w*′*s* parameters constant (Fig. 2), we could solely adjust *a*_0_(*λ*_*i*_) to *a*_0_(*λ*_3_)= 0.002 and *a*_0_(*λ*_4_)= 0.001 respectively while maintaining acceptable accuracy of predictions. The sharp decline in *a*_0_(*λ*_*i*_) values over time reflects the immense effect of the vaccine campaign on the severity of the pandemic and the transition to virus variants being less harmful in terms of severe cases (i.e. deaths). The variability in *a*_0_(*λ*_*i*_) is expected as over the course of the entire pandemic, with yearly average *H*_*R*_ values, city layouts and census data exposed to minimal changes, *w* weights parameters should not vary to a noticeable extent. It is also important to note that the model significantly overestimated *CFR*(*λ*_1_) for 9 cities (Fig. 4a, Table S5) where measured *CFR*(*λ*_1_) were very close to *a*_0_(*λ*_1_). Upon further investigation, these cities were found to be either isolated territories (such as Maldives or Malta, which based on size and population are treated as cities) or quick to adopt social distancing and/or lockdown measures (as in San Francisco CA, Berlin in Germany, or Singapore) during the first wave, *λ*_1_, of the COVID-19 pandemic. We can conclude therefore that measured *CFR* as predicted by the coronavirus severity equation (Eq.2) reflects the absence or inefficiency of sanitary measures taken by local and/or national authorities to manage the first wave of COVID-19.

Multi-spatial scale application of the model with no adjustment to its weight factors allows us to study the impact of specific variables that make up the three distinctive urban factors. Relative humidity takes 90.5% of *O*_*i*_, while *φ* (characterizing city texture, see SI) is responsible for the remaining 9.5%. High percentage value of *H*_*R*_ supports the airborne nature of pandemic transmission underlying the role of evaporation and thermodynamic conditions of the surrounding atmosphere (7, 9). The effect of city texture parameter *φ* can be linked to persistence of active outdoor forms of SARS-CoV-2 previously related to urbanization and particulate atmospheric pollution (13, 23). This also suggests that ordered districts in a given city (i.e. Brooklyn in NYC) are more likely to retain laden droplets than less ordered ones (i.e. Bronx in NYC). Indoor Factor, defined by the household size and density of units have similar contribution, 54% and 46%, respectively, which can be explained by the close-range contaminations and circulation of the virus not just inside units, but also in between them due air fluxes and recirculation (24, 25) and its presence on common surfaces and spaces such as elevators, stair cases, lobbies (8, 26). Since the higher density of inhabitants per housing unit results in higher probability of *CFR*, this work reveals an aspect of social discriminations linked to wealth and living standards. Finally, the Personal Factor is derived solely from population older than 65 years old, which suggests that the current healthcare systems in place across the 30 cities we considered, were able to respond to patient’s needs regardless of their income.

Reduction in global *a*_0_ by a factor 4 (from 0.0089 to 0.0022) during the first three waves followed by a further reduction by 2.8 between the third and fourth waves (from 0.0022 to 0.0008) reflects the joint positive impact of social distancing, hygiene measures and above all vaccination combined with the virus mutations to less harmful variants (i.e. Omicron and subsequent more recent versions of the virus). Each *U*_*i*_ changes predicted *CFR* according to expected norms that is an increase in relative humidity, density of urban dwellers or the proportion of population above 65 years of age, all increase *CFR* values. It is demonstrated that the approach remains operative at the global city scale making model fit predictions with *R*^2^= 0.95 and a low *RMSE* = 0.31 population adjusted *CFR*_*pop*_. It is interesting to note that averaging measured *CFR*s, across all US and UK cities considered in this work, gives comparable to cumulative at country scale values for *CFR*(*λ*_3_): 2.0% vs 1.8% (US) and 2.7% vs. 2.7% (UK) and *CFR*(*λ*_4_): 1.1% vs 1.2% (US) and 0.9% vs. 1.0% (UK) reflecting the fact that the COVID-19 pandemic is mainly an urban phenomenon.

In the field of epidemiology, it is often considered that the *CFR* has limited time validity (usually a few months) and is spatially restricted to a country level. This work demonstrates, however, that the *CFR* carries a multiscale length and time scale stability and thus not only can be used to infer disease severity in both real time and during the entire pandemic. It can also be averaged by location from the zip code to city scales worldwide. Therefore, predicting the severity of COVID-19 using *CFR* provides new dynamic means for evaluating cities in real-time with changing urban factors to equip decision makers and public authorities with quantifiable data to optimize and target their seasonal response to airborne infectious diseases, such as COVID-19 at any geo-spatial scale.

## Materials and Methods

For all input data, we resort to using publicly available building footprints, weather, and census data (Tables S1-S2). We define the first *U*_*i*_, Personal Factor as:

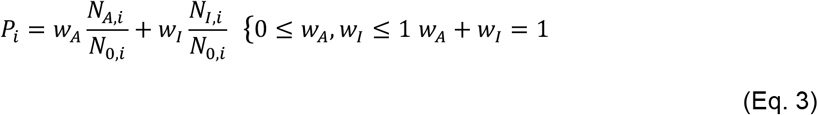

where *w*_*A*_ is a weight factor for inhabitants of age, (*A, i*)≥ 65 years old, *N*_*A,i*_is the total number of people *A, i* ≥ 65, *w*_*I*_ is a weight factor for an annual income, *I, i* ≤ $50,000, *N*_*I,i*_ is the total number of people with *I, i* ≤ $50,000, and *N*_0,*i*_ is the total number of people living in the region *i*. The second *U*_*i*_, Indoor Factor is defined as:

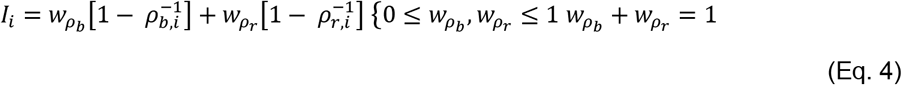

where 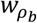 is a weight factor for building density, *ρ*_*b,i*_ (number of housing units per building), and 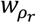 is a weight factor for resident density, *ρ*_*r,i*_ (household size, which is the number of residents per housing unit) in region *i*. The final *U*_*i*_, Outdoor Factor is calculated as:

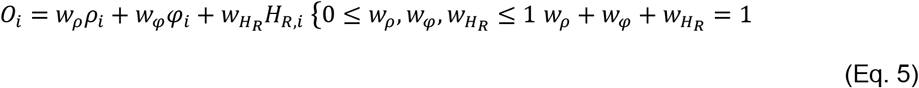

where *w*_*ρ*_ is a weight factor for the planar density of building footprints, *ρ*_*i*_ (ratio between total area of buildings and area of region *i*), *w*_*φ*_ is a weight factor for the order parameter of buildings, *φ*_*i*_, and 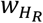 is a weight factor for the relative humidity *H*_*R,i*_, which is assumed constant at the city-scale, except for New York, Los Angeles and Chicago, where city district-level *H*_*R,i*_ was available. The buildup surface area and the local order parameter, *φ*_*i*_ reflects the local urban texture of the first shell of neighboring building with respect to a central one and is used to quantify 2-D angular order of a local urban texture obtained from the 2-D building-building pair correlation function, *g*(*r*)((27), SI), which is defined as:

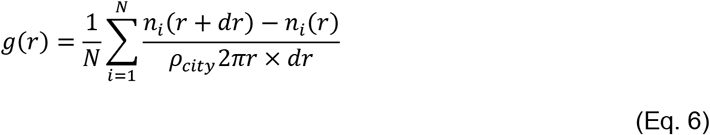

where *n*_*i*_(*r*)*n*_*i*_(*r*) denotes the number of buildings within the radial distance *r* from building *i*, and *dr* is distance increment, which for *g*(*r*) calculations we chose to be 5% of the average building size, *L*. From the distribution of *g*(*r*) we can directly obtain coordination number, *C*_*n*_ (i.e. average number of neighboring buildings) using its integral in the form:

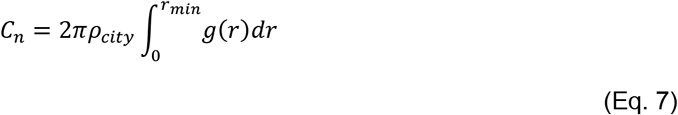

The approach for selecting *C*_*n*_ should be evaluated based on its application, which depending on desired accuracy may lead to different results. Here, the application is defining average local configuration of buildings as captured by *g*(*r*) thus leading to our use of Eq. 7, where *r*_*min*_ is the first local minimum in the *g*(*r*) distribution following the main peak (i.e. one with the maximum *g*(*r*) value). Due to variability in local city texture between zip codes or wards within a given city, integral of the first peak may not always lead to *C*_*n*_ > 1, which is required to be able to obtain angular order parameter with a minimum configuration of 3 buildings, or 2 neighbors. If such situation exists, we proceed with the next local minimum in the distribution of *g*(*r*) until *C*_*n*_ > 1 has been obtained. With such defined *C*_*n*_ we proceed with calculations of the second city texture value, order parameter. Exact *r*_*min*_ values used to define the first peak are presented in Tables S1-S3, with *g*(*r*) functions visualized in figs. S2-S4. Applied at the city scale, we use the absolute values of *C*_*n*_, to find *φ*, which characterizes the average angular distortion of buildings compared to a perfect angular local order of a city at fixed *m* = *C*_*n*_ within the first shell distance determined from the integral of *g*(*r*)(Eq. 7):

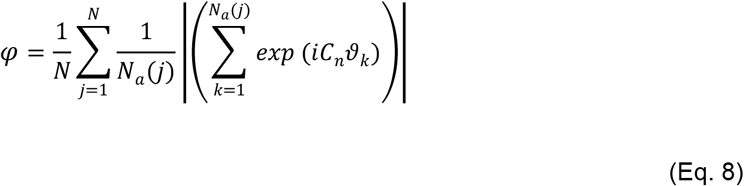

where, *N* is the number of buildings.

The model parameters have been optimized by minimizing the error between predicted and measured *CFR*(*λ*_1_) values using standard statical methods. Although, it is common to apply sum of square errors when quantifying errors in regression analyses, here we adopted a sum of absolute errors, *ε* in the form of:

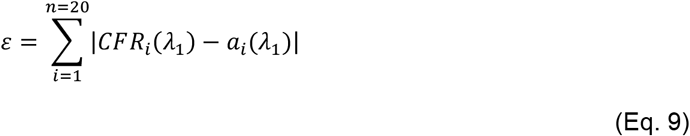

where *i* is the city district ID from Table S4. Since *CFR* values are fractions, differences between any predicted and measured values lead to small fractions and thus squaring the difference would lead to even smaller values thereby introducing bias to any optimization approach that is trying to minimize the sum of errors. As such the absolute sum *ε* provides a more stable optimization parameter. We minimized *ε* using a non-linear generalized reduced gradient (GRD) method with constraint convergence of 0.00001, forward derivates population size of 1000. With such defined parameters, we obtained optimized urban factors and intrinsic *CFR, a*_0_(*λ*_1_)(Fig. 3).

## Data Availability

All data produced in the present work are contained in the manuscript and supplementary information (SI) with url links in SI figures to access publicly available raw data.

## Acknowledgments

We acknowledge CNRS-Innovation for the SURe project funding (www.sure-tech.fr) through the pre-maturation and RISE programs.

## References

1. K. Mouratidis, How COVID-19 reshaped quality of life in cities: A synthesis and implications for urban planning. Land Use Policy 111, 105772 (2021).

2. A. Sharifi, A. R. Khavarian-Garmsir, The COVID-19 pandemic: Impacts on cities and major lessons for urban planning, design, and management. Science of The Total Environment 749, 142391 (2020).

3. M. Dashtbali, M. Mirzaie, A compartmental model that predicts the effect of social distancing and vaccination on controlling COVID-19. Sci Rep 11, 8191 (2021).

4. P. D. Lunn, et al., Motivating social distancing during the COVID-19 pandemic: An online experiment. Social Science & Medicine 265, 113478 (2020).

5. N. Phillips, The coronavirus is here to stay — here’s what that means. Nature 590, 382–384 (2021).

6. C. P. Gerba, “Applied and Theoretical Aspects of Virus Adsorption to Surfaces” in Advances in Applied Microbiology, A. I. Laskin, Ed. (Academic Press, 1984), pp. 133–168.

7. L. Zhao, Y. Qi, P. Luzzatto-Fegiz, Y. Cui, Y. Zhu, COVID-19: Effects of Environmental Conditions on the Propagation of Respiratory Droplets. Nano Lett. 20, 7744–7750 (2020).

8. S. Riddell, S. Goldie, A. Hill, D. Eagles, T. W. Drew, The effect of temperature on persistence of SARS-CoV-2 on common surfaces. Virology Journal 17, 145 (2020).

9. K. A. Prather, et al., Airborne transmission of SARS-CoV-2. Science (2020) (January 30, 2022).

10. L. Bourouiba, Turbulent Gas Clouds and Respiratory Pathogen Emissions: Potential Implications for Reducing Transmission of COVID-19. JAMA 323, 1837–1838 (2020).

11. R. Mittal, R. Ni, J.-H. Seo, The flow physics of COVID-19. Journal of Fluid Mechanics 894 (2020).

12. X. Wu, R. C. Nethery, M. B. Sabath, D. Braun, F. Dominici, Exposure to air pollution and COVID-19 mortality in the United States: A nationwide cross-sectional study. 2020.04.05.20054502 (2020).

13. L. Setti, et al., SARS-Cov-2RNA found on particulate matter of Bergamo in Northern Italy: First evidence. Environ Res 188, 109754 (2020).

14. O. Faye, et al., Use of Viremia to Evaluate the Baseline Case Fatality Ratio of Ebola Virus Disease and Inform Treatment Studies: A Retrospective Cohort Study. PLOS Medicine 12, e1001908 (2015).

15. E. Pujadas, et al., SARS-CoV-2 viral load predicts COVID-19 mortality. The Lancet Respiratory Medicine 8, e70 (2020).

16. A. L. Hill, The math behind epidemics. Physics Today 73, 28–34 (2020).

17. G. Fan, et al., Decreased Case Fatality Rate of COVID-19 in the Second Wave: A study in 53 countries or regions. Transbound Emerg Dis 68, 213–215 (2021).

18. C.-S. Chang, et al., The computation of case fatality rate for novel coronavirus (COVID-19) based on Bayes theorem. Medicine (Baltimore) 99, e19925 (2020).

19. S. Gupta, K. Kumar Patel, S. Sivaraman, A. Mangal, Global Epidemiology of First 90 Days into COVID-19 Pandemic: Disease Incidence, Prevalence, Case Fatality Rate and Their Association with Population Density, Urbanisation and Elderly Population. Journal of Health Management 22, 117–128 (2020).

20. C. S. Narayanan, A novel cohort analysis approach to determining the case fatality rate of COVID-19 and other infectious diseases. PLOS ONE 15, e0233146 (2020).

21. M. Neil, N. Fenton, M. Osman, S. McLachlan, Bayesian network analysis of Covid-19 data reveals higher infection prevalence rates and lower fatality rates than widely reported. Journal of Risk Research 23, 866–879 (2020).

22. J. T. Wu, et al., Estimating clinical severity of COVID-19 from the transmission dynamics in Wuhan, China. Nat Med 26, 506–510 (2020).

23. Y. Yao, et al., Association of particulate matter pollution and case fatality rate of COVID-19 in 49 Chinese cities. Science of The Total Environment 741, 140396 (2020).

24. R. K. Bhagat, M. S. D. Wykes, S. B. Dalziel, P. F. Linden, Effects of ventilation on the indoor spread of COVID-19. Journal of Fluid Mechanics 903 (2020).

25. E. S. Mousavi, N. Kananizadeh, R. A. Martinello, J. D. Sherman, COVID-19 Outbreak and Hospital Air Quality: A Systematic Review of Evidence on Air Filtration and Recirculation. Environ Sci Technol 55, 4134–4147 (2021).

26. N. van Doremalen, et al., Aerosol and Surface Stability of SARS-CoV-2 as Compared with SARS-CoV-1. New England Journal of Medicine 382, 1564–1567 (2020).

27. J. M. Sobstyl, T. Emig, M. J. A. Qomi, F.-J. Ulm, R. J.-M. Pellenq, Role of City Texture in Urban Heat Islands at Nighttime. Phys. Rev. Lett. 120, 108701 (2018).

