## Supplementary Information for "How do Urban Factors Control the Severity of COVID-19?"

* Roland J.M. Pellenq.


| Country | $CFR$ | Country | $CFR$ | Country | $CFR$ | Country | $CFR$ | Country | $CFR$ | Country | $CFR$ | Country | $CFR$ |
|---|---|---|---|---|---|---|---|---|---|---|---|---|---|
| Afghanistan | 0.046 | Cabo Verde | 0.007 | Falkland Islands | 0.000 | Israel | 0.003 | Monaco | 0.006 | Qatar | 0.002 | Sudan | 0.059 |
| Albania | 0.013 | Cambodia | 0.025 | Faroe Islands | 0.001 | Italy | 0.013 | Mongolia | 0.005 | South Korea | 0.008 | Suriname | 0.017 |
| Algeria | 0.026 | Cameroon | 0.016 | Fiji | 0.013 | Jamaica | 0.021 | Montenegro | 0.012 | Moldova | 0.024 | Sweden | 0.007 |
| American Samoa | 0.000 | Canada | 0.011 | Finland | 0.004 | Japan | 0.007 | Montserrat | 0.006 | Réunion | 0.003 | Switzerland | 0.006 |
| Andorra | 0.004 | Cayman Islands | 0.001 | France | 0.007 | Jersey | 0.003 | Morocco | 0.014 | Romania | 0.027 | Syria | 0.058 |
| Angola | 0.019 | Central African Republi | 0.008 | French Guiana | 0.005 | Jordan | 0.011 | Mozambique | 0.010 | Russian Federation | 0.028 | Tajikistan | 0.007 |
| Anguilla | 0.003 | Chad | 0.027 | French Polynesi | 0.013 | Kazakhstan | 0.014 | Myanmar | 0.036 | Rwanda | 0.011 | Thailand | 0.009 |
| Antigua and Barbuda | 0.019 | Chile | 0.019 | Gabon | 0.006 | Kenya | 0.017 | Namibia | 0.025 | Saba | 0.000 | United Kingdom | 0.009 |
| Argentina | 0.015 | China | 0.041 | Gambia | 0.031 | Kiribati | 0.000 | Nauru | 0.000 | Saint Barthelemy | 0.001 | Timor-Leste | 0.006 |
| Armenia | 0.022 | Colombia | 0.023 | Georgia | 0.013 | Kosovo | 0.015 | Nepal | 0.012 | Saint Helena | 0.000 | Togo | 0.007 |
| Aruba | 0.006 | Comoros | 0.020 | Germany | 0.012 | Kuwait | 0.005 | Netherlands | 0.005 | Saint Kitts and Nevis | 0.006 | Tokelau | 0.000 |
| Australia | 0.002 | Congo | 0.015 | Ghana | 0.009 | Kyrgyzstan | 0.015 | New Caledonia | 0.015 | Saint Lucia | 0.016 | Tonga | 0.000 |
| Austria | 0.007 | Congo DRC | 0.015 | Gibraltar | 0.008 | Laos | 0.004 | New Zealand | 0.003 | Saint Martin | 0.004 | Trinidad and Tobago | 0.031 |
| Azerbaijan | 0.013 | Cook Islands | 0.000 | Greece | 0.012 | Latvia | 0.013 | Nicaragua | 0.016 | Saint Pierre & Miquelon | 0.000 | Tunisia | 0.029 |
| Bahamas | 0.023 | Costa Rica | 0.011 | Greenland | 0.000 | Lebanon | 0.010 | Niger | 0.034 | Saint Vincent & the Grenadine | 0.012 | Turkey | 0.008 |
| Bahrain | 0.004 | Côte d'Ivoire | 0.010 | Grenada | 0.017 | Lesotho | 0.022 | Nigeria | 0.012 | Samoa | 0.000 | Turkmenistan | 0.000 |
| Bangladesh | 0.016 | Croatia | 0.015 | Guadeloupe | 0.008 | Liberia | 0.040 | Niue | 0.000 | San Marino | 0.008 | Turks & Caicos Island | 0.006 |
| Barbados | 0.006 | Cuba | 0.008 | Guam | 0.011 | Libya | 0.014 | North Macedonia | 0.031 | Sao Tome and Principe | 0.012 | Tuvalu | 0.000 |
| Belarus | 0.008 | Curacao | 0.006 | Guatemala | 0.024 | Liechtenstein | 0.008 | Northern Mariana Island | 0.005 | Saudi Arabia | 0.013 | Uganda | 0.022 |
| Belgium | 0.009 | Cyprus | 0.003 | Guernsey | 0.003 | Lithuania | 0.012 | Norway | 0.002 | Senegal | 0.023 | Ukraine | 0.025 |
| Belize | 0.012 | Czech Republic | 0.012 | Guinea | 0.012 | Luxembourg | 0.006 | Palestinian Territory | 0.010 | Serbia | 0.008 | United Arab Emirates | 0.003 |
| Benin | 0.006 | North Korea | 0.000 | Guinea-Bissau | 0.021 | Madagascar | 0.021 | Oman | 0.012 | Seychelles | 0.004 | Tanzania | 0.024 |
| Bermuda | 0.011 | Denmark | 0.002 | Guyana | 0.020 | Malawi | 0.030 | Other | 0.017 | Sierra Leone | 0.016 | United States | 0.012 |
| Bhutan | 0.001 | Djibouti | 0.012 | Haiti | 0.027 | Malaysia | 0.011 | Pakistan | 0.021 | Singapore | 0.002 | US Virgin Islands | 0.007 |
| Bolivia | 0.025 | Dominica | 0.006 | Holy See | 0.000 | Maldives | 0.002 | Palau | 0.000 | Sint Eustatius | 0.003 | Uruguay | 0.010 |
| Bonaire | 0.004 | Dominican Republic | 0.008 | Honduras | 0.027 | Mali | 0.024 | Panama | 0.011 | Sint Maarten | 0.009 | Uzbekistan | 0.007 |
| Bosnia and Herzegovin | 0.041 | Ecuador | 0.048 | Hungary | 0.027 | Malta | 0.008 | Papua New Guinea | 0.016 | Slovakia | 0.018 | Vanuatu | 0.000 |
| Botswana | 0.010 | Egypt | 0.053 | Iceland | 0.001 | Marshall Islands | 0.000 | Paraguay | 0.030 | Slovenia | 0.009 | Venezuela | 0.011 |
| Brazil | 0.025 | El Salvador | 0.029 | India | 0.012 | Martinique | 0.009 | Peru | 0.064 | Solomon Islands | 0.008 | Vietnam | 0.017 |
| British Virgin Islands | 0.008 | Equatorial Guinea | 0.012 | Indonesia | 0.033 | Mauritania | 0.016 | Philippines | 0.015 | Somalia | 0.053 | Wallis & Futuna | 0.015 |
| Brunei Darussalam | 0.004 | Eritrea | 0.010 | Iran | 0.021 | Mauritius | 0.011 | Pitcairn | 0.000 | South Africa | 0.026 | Yemen | 0.183 |
| Bulgaria | 0.035 | Estonia | 0.006 | Iraq | 0.011 | Mayotte | 0.005 | Poland | 0.022 | South Sudan | 0.008 | Zambia | 0.013 |
| Burkina Faso | 0.018 | Eswatini | 0.020 | Ireland | 0.005 | Mexico | 0.062 | Portugal | 0.008 | Spain | 0.009 | Zimbabwe | 0.023 |
| Burundi | 0.000 | Ethiopia | 0.016 | Isle of Man | 0.003 | Micronesia | 0.000 | Puerto Rico | 0.008 | Sri Lanka | 0.025 | | |

**Table S1.** Cumulative $CFR$ data for countries for period between March 2020 and January 2022 obtained from World Health Organization (WHO).

The objective of this work is to provide a model that provides accurate predictions of the severity of COVID-19—captured with the Case Fatality Ratio $CFR$ (Eq.1, Fig. 2a)—by using urban factors derived using Eqs.3-5. Input data for this work is publicly available and has been obtained from census, weather and building footprints repositories (Table S2, Table S3). $CFR$ is a critical and important measure of the severity of a disease/virus, as it captures mortality of population with varied social profiles and habits, including age, health, population density etc. as well as environmental conditions such as temperature, relative humidity, atmospheric pollution, or UV radiation. Therefore, for the same variant of COVID-19 disease, the $CFR$ can vary between different geographical locations, including cities or countries (Fig. 1, Table S1). The accurate determination of the $CFR$ has been the topic of many scientific studies and is considered to be a key quantity for targeted emergency and health care infrastructure deployment (1–6). Being a ratio of deaths and confirmed cases of COVID-19, $CFR$ thus is only as accurate as the testing methods to report the number of cases and deaths. At the beginning of the corona virus pandemic spread, there were very limited capabilities to precisely evaluate at the scale of countries the number of infected people with corresponding mortality rates; more importantly, however, these values were still on the rise, which based on the distribution analysis of the COVID-19 data (Figs. 2a-b, S2) appear to have become more stable six to eight months after the beginning of the pandemic (March 2020) signifying the end of the first wave, $\lambda_1$. Thus, $\lambda_1$ period was selected for the evaluation of $CFR$. Although, other waves of the pandemic should also be studied to understand the underlying factors attributing to variations in $CFR$ for the same geographical location, due to limitations of COVID-19 data (confirmed cases and deaths) at the city level and city district levels, in this study the focus is on the first wave, $\lambda_1$ (which for the purposes of this study, depending on the location includes some parts of second wave data between November 2021 and February 2022) and cumulative $CFR$ values for the first, second and third, $\lambda_3$ (March 2020 – June 2021) and the fourth, $\lambda_4$ (March 2020 – January 2022) waves of the COVID-19 pandemic. For any COVID-19 data used in the study, the assumption is that all reported data for any given location is for its residents only.

| ID | City | Country | COVID-19 | Census | Building Footprints | Region Outlines | Humidity |
|---|---|---|---|---|---|---|---|
| 1 | Los Angeles, CA | US | http://publichealth.lacounty.gov/media/Coronavirus/locations.htm | https://censusreporter.org/ | https://planning.lacounty.gov/gis/data | https://data.lacounty.gov/Geospatial/ZIP-Codes/65v5-jw9f | Table S2 |
| 2 | New York, NY | US | https://www1.nyc.gov/site/doh/covid/covid-19-data-totals.page | " | https://www1.nyc.gov/site/doitt/residents/gis-2d-data.page | https://data.cityofnewyork.us/City-Government/Borough-Boundaries/tqmj-j8zm | " |
| 3 | Washington, D.C. | US | https://opendata.dc.gov/datasets/DCGIS::dc-covid-19-cases-by-ward/about | " | https://opendata.dc.gov/datasets/building-footprints | https://opendata.dc.gov/datasets/DCGIS::ward/about | https://weather-and-climate.com, https://en.climate-data.org |
| 4 | Chicago, IL | US | https://data.cityofchicago.org/Health-Human-Services/COVID-19-Cases-Tests-and-Deaths-by-ZIP-Code/yhhz-zm2v | " | https://data.cityofchicago.org/Buildings/Building-Footprints-current-/hz9b-7nh8 | | Table S2 |
| 5 | Seattle, WA | US | https://kingcounty.gov/depts/health/covid-19/data/daily-summary.aspx | " | https://data.seattlecitygis.opendata.arcgis.com/datasets/building-outlines-2015 | https://data-seattlecitygis.opendata.arcgis.com/datasets/zip-codes | " |
| 6 | Boston, MA | US | https://www.boston.gov/news/coronavirus-disease-covid-19-boston | " | https://bostonopendata-boston.opendata.arcgis.com/datasets/boston-buildings | not applicable | https://en.climate-data.org, https://weather-and-climate.com, https://www.weather-us.com |
| 7 | Denver, CO | US | https://www.denvergov.org/Government/COVID-19-Information/COVID-19-in-Denver/COVID-19-Case-Dashboards | " | https://www.denvergov.org/opendata/dataset/city-and-county-of-denver-building-outlines-2016 | not applicable | " |
| 8 * | San Francisco, CA | US | https://sf.gov/data/covid-19-cases-and-deaths | " | https://data.sfgov.org/Geographic-Locations-and-Boundaries/Building-Footprints/ynuv-fyni | not applicable | " |
| 9 | Philadelphia, PA | US | https://www.health.pa.gov/topics/disease/coronavirus/Pages/Coronavirus.aspx | " | https://www.opendataphilly.org/dataset/buildings | not applicable | " |
| 10 | London | UK | https://data.london.gov.uk/dataset/coronavirus--covid-19--cases | https://data.london.gov.uk/census/data/ | http://download.geofabrik.de/ | https://osm-boundaries.com/Map | https://en.climate-data.org, https://weather-and-climate.com, https://www.weather-atlas.com |
| 11 | Glasgow | UK | https://coronavirus.data.gov.uk/ | https://www.nrscotland.gov.uk/files/statistics/council-area-data-sheets/glasgow-city-council-profile.html | " | " | " |
| 12 | Aberdeen | UK | " | https://www.nrscotland.gov.uk/files/statistics/council-area-data-sheets/aberdeen-city-council-profile.html | " | " | " |
| 13 | Bristol | UK | " | https://opendata.bristol.gov.uk/pages/homepage/ | " | " | https://en.climate-data.org, https://www.weather-atlas.com |
| 14 | Edinburgh | UK | " | https://www.nrscotland.gov.uk/files/statistics/council-area-data-sheets/city-of-edinburgh-council-profile.html | " | " | https://en.climate-data.org, https://weather-and-climate.com, https://www.weather-atlas.com |
| 15 * | Berlin | Germany | https://www.berlin.de/corona/lagebericht/#barrierefreiheit | https://www.citypopulation.de/ | " | " | " |
| 16 | Balearic Islands | Spain | https://www.sanidad.gob.es/profesionales/saludPublica/ccayes/alertasActual/nCov/situacionActual.htm | " | " | " | https://weather-and-climate.com |
| 17 | Madrid | Spain | https://www.comunidad.madrid/servicios/salud/coronavirus#situacion-epidemiologica-actual | " | " | " | https://en.climate-data.org, https://weather-and-climate.com |
| 18 | Rio De Janeiro | Brazil | https://covid.saude.gov.br/ | " | " | " | https://en.climate-data.org, https://weather-and-climate.com, https://www.weather-atlas.com |
| 19 | São Paulo | Brazil | " | " | " | " | " |
| 20 | Recife | Brazil | " | " | " | " | " |
| 21 | Porto Alegre | Brazil | " | " | " | " | " |
| 22 | Barranquilla | Colombia | http://www.ins.gov.co/Noticias/Paginas/Coronavirus.aspx | " | " | " | https://en.climate-data.org, https://weather-and-climate.com |
| 23 * | Chennai | India | https://www.mygov.in/covid-19 | " | " | " | https://en.climate-data.org, https://weather-and-climate.com, https://www.weather-atlas.com |
| 24 * | Delhi | India | " | " | " | " | " |
| 25 | Kokata | India | " | " | " | " | " |
| 26 * | n/a | Bahrain | https://www.worldometers.info/coronavirus/ | https://data.worldbank.org/country/bahrain | " | " | https://weather-and-climate.com |
| 27 * | n/a | Liechtenstein | " | https://countrymeters.info/en/Liechtenstein | " | " | " |
| 28 * | n/a | Maldives | " | https://data.worldbank.org/country/maldives | " | " | " |
| 29 * | n/a | Malta | " | https://data.worldbank.org/country/malta | " | " | " |
| 30 * | Singapore | Singapore | " | https://data.worldbank.org/country/singapore | " | " | https://en.climate-data.org, https://weather-and-climate.com, https://www.weather-atlas.com |
| P ** | Paris | France | not available | https://www.citypopulation.de/ | " | " | " |
| M ** | Marseille | France | " | " | " | " | " |
| L ** | Lyon | France | " | " | " | " | " |

**Table S2.** Sources for data for city districts and cities. Multiple sources for humidity were used to obtain average values presented in Table S5.

| City | ID | ID-SI | District | Humidty Source | Location |
|---|---|---|---|---|---|
| New York NY | 1 | 1 | Bronx | https://forecast.weather.gov | La Guardia Airport, NY |
| | 2 | 2 | Brooklyn | " | JFK Airport, NY |
| | 3 | 3 | Manhattan | " | Central Park, NY |
| | 4 | 4 | Queens | " | JFK Airport, NY |
| | 5 | 5 | Staten Island | " | Newark, NJ |
| Los Angeles CA | 6 | 1 | North | https://www.titudorancea.com | Van Nuys, CA |
| | 7 | 2 | West | https://www.laalmanac.com | LAX Airport, CA |
| | 8 | 3 | Central | https://www.titudorancea.com | USC Campus, LA Downtown, CA |
| | 9 | 4 | North East | " | Burbank, CA |
| | 10 | 5 | East | " | Riverside, CA |
| | 11 | 6 | South | " | Long Beach, CA |
| | 12 | 7 | South East | " | Fullerton, CA |
| Chicago IL | 13 | 1 | North | https://weather-and-climate.com, https://en.climate-data.org | Chicago IL |
| | 14 | 2 | Central East | https://www.myweather2.com | Midway Airport, IL |
| | 15 | 3 | Central West | https://weather-and-climate.com, https://en.climate-data.org | Chicago IL |
| | 16 | 4 | Central South | https://www.myweather2.com | Midway Airport, IL |
| | 17 | 5 | South | https://weather-and-climate.com, https://en.climate-data.org | Chicago IL |
| Seattle WA | 18 | 1 | North | https://weather-and-climate.com, https://en.climate-data.org | Seattle WA |
| | 19 | 2 | Central | " | " |
| | 20 | 3 | South | " | " |

**Table S3.** Sources for humidity data for city districts data used for model optimization (Fig. 4a). All sources for each location were used to derive humidity values in Table S4.

| City | ID | ID-SI | District | $CFR_s(\lambda_1)$ | $CFR(\lambda_1)$ | $CFR_m(\lambda_1)$ | $N_{a,i}/N_{0,i}$ | $N_{I,i}/N_{0,i}$ | $P_i$ | $B$ | $B_a$ | $B_u$ | $P$ | $\rho_{r,i}$ | $\rho_{b,i}$ | $I_i$ | $R_{lim}(m)$ | $C_{n,1}$ | $\varphi$ | $\rho_i$ | $H_{R,i}$ | $O_i$ | $a_i(\lambda_1)$ |
|---|---|---|---|---|---|---|---|---|---|---|---|---|---|---|---|---|---|---|---|---|---|---|---|
| New York NY | 1 | 1 | Bronx | 0.095 | 0.078 | 0.078 | 0.130 | 0.56 | 0.130 | 105,194 | 102,514 | 537,197 | 1,418,207 | 2.64 | 5.24 | 0.708 | 13.8 | 2.64 | 0.515 | 0.193 | 0.58 | 0.576 | 0.0818 |
| | 2 | 2 | Brooklyn | 0.126 | 0.092 | 0.093 | 0.140 | 0.40 | 0.140 | 332,844 | 317,589 | 1,065,363 | 2,559,903 | 2.40 | 3.35 | 0.638 | 9.7 | 2.17 | 0.804 | 0.267 | 0.67 | 0.689 | 0.0924 |
| | 3 | 3 | Manhattan | 0.142 | 0.088 | 0.091 | 0.170 | 0.31 | 0.170 | 46,210 | 45,583 | 892,930 | 1,628,706 | 1.82 | 19.59 | 0.681 | 20.8 | 4.12 | 0.554 | 0.327 | 0.63 | 0.625 | 0.0884 |
| | 4 | 4 | Queens | 0.107 | 0.080 | 0.081 | 0.160 | 0.34 | 0.160 | 462,393 | 441,458 | 869,346 | 2,253,858 | 2.59 | 1.97 | 0.558 | 10.3 | 2.26 | 0.764 | 0.196 | 0.67 | 0.686 | 0.0766 |
| | 5 | 5 | Staten Island | 0.051 | 0.057 | 0.059 | 0.170 | 0.29 | 0.170 | 141,787 | 139,557 | 181,765 | 476,143 | 2.62 | 1.30 | 0.440 | 20.5 | 3.88 | 0.595 | 0.114 | 0.60 | 0.600 | 0.0478 |
| Los Angeles CA | 6 | 1 | North | 0.067 | 0.053 | 0.054 | 0.126 | 0.35 | 0.126 | 494,156 | 441,080 | 585,322 | 1,630,595 | 2.79 | 1.33 | 0.459 | 27.1 | 4.50 | 0.497 | 0.175 | 0.52 | 0.522 | 0.0393 |
| | 7 | 2 | West | 0.098 | 0.060 | 0.061 | 0.158 | 0.29 | 0.158 | 201,718 | 188,483 | 341,485 | 659,757 | 1.93 | 1.81 | 0.467 | 27.1 | 5.28 | 0.443 | 0.166 | 0.69 | 0.672 | 0.0593 |
| | 8 | 3 | Central | 0.065 | 0.052 | 0.052 | 0.115 | 0.52 | 0.115 | 326,335 | 300,166 | 569,214 | 1,456,177 | 2.56 | 1.90 | 0.546 | 23.0 | 4.73 | 0.456 | 0.250 | 0.62 | 0.610 | 0.0562 |
| | 9 | 4 | North East | 0.060 | 0.054 | 0.055 | 0.153 | 0.41 | 0.153 | 262,017 | 254,951 | 260,517 | 812,164 | 3.12 | 1.02 | 0.376 | 26.7 | 4.76 | 0.456 | 0.221 | 0.54 | 0.538 | 0.0363 |
| | 10 | 5 | East | 0.019 | 0.040 | 0.042 | 0.132 | 0.31 | 0.132 | 189,240 | 188,187 | 236,349 | 825,003 | 3.49 | 1.26 | 0.478 | 28.8 | 3.47 | 0.445 | 0.142 | 0.53 | 0.528 | 0.0414 |
| | 11 | 6 | South | 0.050 | 0.048 | 0.051 | 0.131 | 0.39 | 0.131 | 477,080 | 458,804 | 517,343 | 1,503,181 | 2.91 | 1.13 | 0.406 | 27.5 | 5.37 | 0.455 | 0.211 | 0.66 | 0.645 | 0.0479 |
| | 12 | 7 | South East | 0.042 | 0.045 | 0.046 | 0.125 | 0.39 | 0.125 | 364,732 | 362,879 | 365,877 | 1,274,096 | 3.48 | 1.01 | 0.388 | 27.3 | 4.76 | 0.457 | 0.229 | 0.61 | 0.597 | 0.0409 |
| Chicago IL | 13 | 1 | North | 0.056 | 0.050 | 0.050 | 0.120 | 0.37 | 0.120 | 192,204 | 189,474 | 227,553 | 533,020 | 2.34 | 1.20 | 0.386 | 12.2 | 2.06 | 0.896 | 0.206 | 0.75 | 0.762 | 0.0701 |
| | 14 | 2 | Central East | 0.044 | 0.044 | 0.044 | 0.100 | 0.52 | 0.100 | 137,285 | 136,160 | 195,261 | 500,556 | 2.56 | 1.43 | 0.468 | 12.2 | 2.12 | 0.871 | 0.250 | 0.65 | 0.669 | 0.0569 |
| | 15 | 3 | Central West | 0.119 | 0.101 | 0.106 | 0.070 | 0.35 | 0.070 | 21,111 | 20,942 | 62,737 | 126,151 | 2.01 | 3.00 | 0.578 | 10.9 | 2.10 | 0.858 | 0.266 | 0.75 | 0.760 | 0.0947 |
| | 16 | 4 | Central South | 0.035 | 0.035 | 0.043 | 0.130 | 0.56 | 0.130 | 176,658 | 174,306 | 153,855 | 420,983 | 2.74 | 0.88 | 0.281 | 12.9 | 2.11 | 0.869 | 0.207 | 0.65 | 0.669 | 0.0436 |
| | 17 | 5 | South | 0.075 | 0.069 | 0.070 | 0.160 | 0.58 | 0.160 | 166,321 | 164,130 | 145,794 | 325,612 | 2.23 | 0.89 | 0.240 | 13.5 | 2.12 | 0.844 | 0.150 | 0.75 | 0.759 | 0.0581 |
| Seattle WA | 18 | 1 | North | 0.106 | 0.089 | 0.092 | 0.130 | 0.28 | 0.130 | 216,962 | 153,887 | 164,909 | 368,281 | 2.23 | 1.07 | 0.328 | 10.8 | 2.67 | 0.530 | 0.183 | 0.76 | 0.753 | 0.0602 |
| | 19 | 2 | Central | 0.079 | 0.066 | 0.067 | 0.120 | 0.28 | 0.120 | 95,087 | 68,308 | 142,618 | 251,008 | 1.76 | 2.09 | 0.473 | 10.9 | 2.66 | 0.544 | 0.208 | 0.76 | 0.754 | 0.0761 |
| | 20 | 3 | South | 0.038 | 0.037 | 0.040 | 0.130 | 0.33 | 0.130 | 190,633 | 128,256 | 105,848 | 255,257 | 2.41 | 0.83 | 0.218 | 11.9 | 2.79 | 0.541 | 0.154 | 0.76 | 0.754 | 0.0520 |

**Table S4.** City Districts measured and predicted $CFR$ data used to derive optimal Urban Factors with their input values. District boundaries are defined using boundaries as shown in Figure S1.

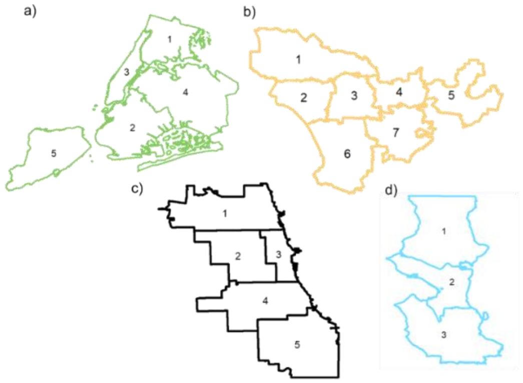


**Fig S1:** (a) New York NY boroughs, (b) Los Angeles CA city and county districts, (c) Chicago IL districts, (d) Seattle WA districts.

| ID | City | Country | $CFR_s(\lambda_1)$ | $CFR(\lambda_1)$ | $CFR_m(\lambda_1)$ | $CFR(\lambda_3)$ | $CFR(\lambda_4)$ | $N_{a,i}/N_{0,i}$ | $P_i$ | $B$ | $B_a$ | $B_u$ | $P$ | $\rho_{r,i}$ | $\rho_{b,i}$ | $I_i$ | $R_{lim}(m)$ | $C_{n,1}$ | $\varphi$ | $\rho_i$ | $H_{R,i}$ | $O_i$ | $a_i(\lambda_1)$ | $a_i(\lambda_3)$ | $a_i(\lambda_4)$ |
|---|---|---|---|---|---|---|---|---|---|---|---|---|---|---|---|---|---|---|---|---|---|---|---|---|---|
| 1 | Los Angeles, CA | US | 0.050 | 0.046 | 0.046 | 0.020 | 0.011 | 0.131 | 0.131 | 2,315,278 | 2,194,550 | 2,643,463 | 8,160,973 | 3.09 | 1.20 | 0.443 | 27.3 | 5.04 | 0.449 | 0.196 | 0.60 | 0.635 | 0.050 | 0.012 | 0.004 |
| 2 | New York, NY | US | 0.109 | 0.087 | 0.088 | 0.035 | 0.017 | 0.151 | 0.151 | 1,088,428 | 1,046,701 | 3,546,601 | 8,336,817 | 2.35 | 3.39 | 0.635 | 9.9 | 2.22 | 0.791 | 0.206 | 0.63 | 0.642 | 0.081 | 0.019 | 0.007 |
| 3 | Washington, D.C. | US | 0.057 | 0.054 | 0.055 | 0.023 | 0.010 | 0.120 | 0.120 | 163,336 | 150,432 | 290,533 | 705,749 | 2.43 | 1.93 | 0.539 | 28.8 | 7.50 | 0.378 | 0.166 | 0.65 | 0.704 | 0.072 | 0.017 | 0.006 |
| 4 | Chicago, IL | US | 0.052 | 0.049 | 0.050 | 0.020 | 0.013 | 0.130 | 0.130 | 801,434 | 685,012 | 1,218,078 | 2,693,959 | 2.21 | 1.78 | 0.497 | 12.5 | 2.08 | 0.875 | 0.200 | 0.71 | 0.713 | 0.072 | 0.017 | 0.006 |
| 5 | Seattle, WA | US | 0.064 | 0.061 | 0.063 | 0.017 | 0.007 | 0.127 | 0.127 | 526,524 | 350,451 | 372,011 | 753,655 | 2.03 | 1.06 | 0.300 | 11.4 | 2.73 | 0.542 | 0.177 | 0.76 | 0.755 | 0.058 | 0.014 | 0.005 |
| 6 | Boston, MA | US | 0.069 | 0.052 | 0.053 | 0.020 | 0.010 | 0.120 | 0.120 | 120,432 | 106,841 | 303,791 | 694,295 | 2.29 | 2.84 | 0.602 | 23.0 | 4.04 | 0.547 | 0.188 | 0.70 | 0.701 | 0.084 | 0.020 | 0.007 |
| 7 | Denver, CO | US | 0.063 | 0.057 | 0.061 | 0.011 | 0.007 | 0.120 | 0.120 | 318,294 | 261,885 | 338,346 | 727,211 | 2.15 | 1.29 | 0.392 | 28.8 | 5.83 | 0.406 | 0.164 | 0.51 | 0.496 | 0.033 | 0.008 | 0.003 |
| 8 * | San Francisco, CA | US | 0.021 | 0.019 | 0.021 | 0.015 | 0.007 | 0.160 | 0.160 | 176,366 | 166,528 | 406,399 | 881,549 | 2.17 | 2.44 | 0.563 | 13.3 | 2.16 | 0.534 | 0.266 | 0.74 | 0.716 | 0.084 | 0.020 | 0.007 |
| 9 | Philadelphia, PA | US | 0.078 | 0.064 | 0.064 | 0.024 | 0.016 | 0.140 | 0.140 | 543,331 | 535,907 | 691,653 | 1,584,064 | 2.29 | 1.29 | 0.407 | 12.4 | 3.35 | 0.522 | 0.199 | 0.69 | 0.682 | 0.054 | 0.013 | 0.004 |
| 10 | London | UK | 0.156 | 0.131 | 0.136 | 0.020 | 0.008 | 0.112 | 0.112 | 646,856 | 646,802 | 3,249,970 | 8,599,250 | 2.65 | 5.02 | 0.705 | 24.1 | 2.34 | 0.737 | 0.144 | 0.77 | 0.786 | 0.157 | 0.037 | 0.013 |
| 11 | Glasgow | UK | 0.164 | 0.168 | 0.174 | 0.030 | 0.012 | 0.140 | 0.140 | 59,346 | 59,339 | 294,622 | 633,120 | 2.15 | 4.97 | 0.656 | 53.0 | 7.95 | 0.392 | 0.118 | 0.83 | 0.816 | 0.155 | 0.036 | 0.013 |
| 12 | Aberdeen | UK | 0.105 | 0.094 | 0.095 | 0.035 | 0.009 | 0.160 | 0.160 | 36,551 | 36,549 | 108,381 | 228,670 | 2.11 | 2.97 | 0.589 | 26.1 | 2.56 | 0.448 | 0.059 | 0.83 | 0.791 | 0.118 | 0.028 | 0.010 |
| 13 | Bristol | UK | 0.205 | 0.212 | 0.214 | 0.019 | 0.006 | 0.130 | 0.130 | 52,802 | 52,765 | 196,100 | 467,099 | 2.38 | 3.72 | 0.650 | 26.8 | 2.59 | 0.621 | 0.150 | 0.82 | 0.809 | 0.148 | 0.035 | 0.012 |
| 14 | Edinburgh | UK | 0.174 | 0.152 | 0.154 | 0.031 | 0.009 | 0.150 | 0.150 | 55,477 | 55,474 | 238,269 | 524,930 | 2.20 | 4.30 | 0.648 | 28.0 | 2.56 | 0.445 | 0.070 | 0.82 | 0.804 | 0.145 | 0.034 | 0.012 |
| 15 * | Berlin | Germany | 0.066 | 0.029 | 0.029 | 0.020 | 0.007 | 0.170 | 0.170 | 98,123 | 88,811 | 2,121,420 | 3,769,495 | 1.78 | 23.89 | 0.677 | 29.1 | 17.10 | 0.432 | 0.038 | 0.73 | 0.728 | 0.118 | 0.028 | 0.010 |
| 16 | Balearic Islands | Spain | 0.173 | 0.102 | 0.105 | 0.014 | n/a | 0.155 | 0.155 | 168,673 | 152,463 | 530,110 | 1,171,543 | 2.21 | 3.48 | 0.624 | 20.5 | 6.64 | 0.410 | 0.008 | 0.72 | 0.698 | 0.090 | 0.021 | 0.007 |
| 17 | Madrid | Spain | 0.126 | 0.133 | 0.141 | 0.021 | 0.018 | 0.204 | 0.204 | 115,122 | 113,309 | 1,318,075 | 3,334,730 | 2.53 | 11.63 | 0.747 | 23.7 | 4.91 | 0.534 | 0.080 | 0.58 | 0.544 | 0.096 | 0.023 | 0.008 |
| 18 | Rio De Janeiro | Brazil | 0.109 | 0.121 | 0.147 | 0.058 | 0.043 | 0.105 | 0.105 | 257,363 | 221,008 | 1,761,832 | 6,747,815 | 3.83 | 7.92 (1.59) | 0.801 | 14.4 | 4.55 | 0.470 | 0.033 | 0.79 | 0.771 | 0.097 | 0.023 | 0.008 |
| 19 | São Paulo | Brazil | 0.083 | 0.078 | 0.086 | 0.034 | 0.040 | 0.081 | 0.081 | 2,169,757 | 1,963,826 | 3,120,312 | 12,325,232 | 3.95 | 1.59 | 0.573 | 18.9 | 7.30 | 0.365 | 0.034 | 0.79 | 0.760 | 0.091 | 0.021 | 0.008 |
| 20 | Recife | Brazil | 0.131 | 0.086 | 0.110 | n/a | n/a | 0.081 | 0.081 | 89,612 | 84,528 | 453,003 | 1,653,461 | 3.65 | 5.36 (1.59) | 0.766 | 18.0 | 5.08 | 0.434 | 0.067 | 0.79 | 0.756 | 0.088 | 0.021 | 0.007 |
| 21 | Porto Alegre | Brazil | 0.076 | 0.042 | 0.058 | n/a | n/a | 0.105 | 0.105 | 489,796 | 435,858 | 396,867 | 1,488,252 | 3.75 | 0.91 | 0.350 | 18.0 | 5.61 | 0.423 | 0.112 | 0.77 | 0.746 | 0.058 | 0.014 | 0.005 |
| 22 | Barranquilla | Colombia | 0.072 | 0.077 | 0.085 | n/a | n/a | 0.097 | 0.097 | 246,843 | 235,235 | 243,985 | 1,273,600 | 5.22 | 1.04 | 0.452 | 15.4 | 4.18 | 0.524 | 0.193 | 0.82 | 0.799 | 0.087 | 0.020 | 0.007 |
| 23 * | Chennai | India | 0.019 | 0.021 | 0.022 | n/a | n/a | 0.095 | 0.095 | 348,054 | 347,707 | 2,025,143 | 7,088,000 | 3.50 | 5.82 | 0.767 | 16.8 | 2.74 | 0.608 | 0.126 | 0.71 | 0.706 | 0.142 | 0.033 | 0.012 |
| 24 * | Delhi | India | 0.038 | 0.034 | 0.041 | n/a | n/a | 0.061 | 0.061 | 523,813 | 521,388 | 3,340,538 | 16,349,831 | 4.89 | 6.41 | 0.818 | 22.6 | 4.42 | 0.416 | 0.067 | 0.47 | 0.479 | 0.099 | 0.023 | 0.008 |
| 25 | Kokata | India | 0.076 | 0.125 | 0.137 | n/a | n/a | 0.077 | 0.077 | 201,625 | 200,817 | 999,265 | 4,496,694 | 4.50 | 4.98 | 0.788 | 24.4 | 4.85 | 0.438 | 0.173 | 0.71 | 0.701 | 0.148 | 0.035 | 0.012 |
| 26 * | n/a | Bahrain | 0.044 | 0.004 | 0.010 | 0.005 | 0.004 | 0.034 | 0.034 | 12,793 | 12,699 | 254,514 | 1,501,635 | 5.90 | 20.04 | 0.886 | 25.5 | 3.06 | 0.361 | 0.012 | 0.73 | 0.703 | 0.262 | 0.061 | 0.022 |
| 27 * | n/a | Liechtensteir | 0.005 | 0.012 | 0.013 | 0.019 | 0.008 | 0.189 | 0.189 | 14,036 | 13,363 | 16,983 | 39,062 | 2.30 | 1.27 | 0.403 | 30.4 | 3.27 | 0.514 | 0.020 | 0.73 | 0.714 | 0.063 | 0.015 | 0.005 |
| 28 * | n/a | Maldives | 0.035 | 0.005 | 0.005 | 0.003 | 0.002 | 0.048 | 0.048 | 18,231 | 17,865 | 77,321 | 402,071 | 5.20 | 4.33 | 0.790 | 32.8 | 5.79 | 0.503 | 0.012 | 0.79 | 0.769 | 0.187 | 0.044 | 0.016 |
| 29 * | n/a | Malta | 0.018 | 0.014 | 0.015 | 0.014 | 0.008 | 0.213 | 0.213 | 17,146 | 16,471 | 190,579 | 514,564 | 2.70 | 11.57 | 0.761 | 39.4 | 5.34 | 0.533 | 0.029 | 0.76 | 0.744 | 0.179 | 0.042 | 0.015 |
| 30 * | Singapore | Singapore | 0.001 | 0.004 | 0.005 | 0.001 | 0.002 | 0.109 | 0.109 | 108,849 | 108,495 | 1,722,972 | 5,685,807 | 3.30 | 15.88 | 0.808 | 16.7 | 3.35 | 0.911 | 0.121 | 0.84 | 0.846 | 0.338 | 0.079 | 0.028 |
| P ** | Paris | France | n/a | n/a | n/a | n/a | n/a | 0.167 | 0.167 | 103,129 | 90,721 | 1,163,423 | 2,175,601 | 1.87 | 12.82 | 0.676 | 26.9 | 5.96 | 0.420 | 0.317 | 0.77 | 0.743 | 0.123 | 0.029 | 0.010 |
| M ** | Marseille | France | n/a | n/a | n/a | n/a | n/a | 0.190 | 0.190 | 288,312 | 170,851 | 377,512 | 868,277 | 2.30 | 2.21 | 0.557 | 13.0 | 4.80 | 0.464 | 0.227 | 0.69 | 0.677 | 0.075 | 0.018 | 0.006 |
| L ** | Lyon | France | n/a | n/a | n/a | n/a | n/a | 0.148 | 0.148 | 46,474 | 38,266 | 259,318 | 518,635 | 2.00 | 6.78 | 0.663 | 27.0 | 6.46 | 0.397 | 0.234 | 0.75 | 0.723 | 0.108 | 0.025 | 0.009 |

**Table S5.** Cities measured and predicted $CFR$ data with Urban Factors and input values used to derive them. *Outliers in $CFR$ predictions – cities with $CFR$ values close to intrinsic $CFR$, $a_0$. **Cities with no availability of $CFR$ due to lack of publicly available COVID-19 raw data (confirmed cases and deaths). Values in () for $\rho_b$ are corrected for the discrepancies in building footprints data due to an incorrect number of buildings. n/a shows lack of publicly available data.

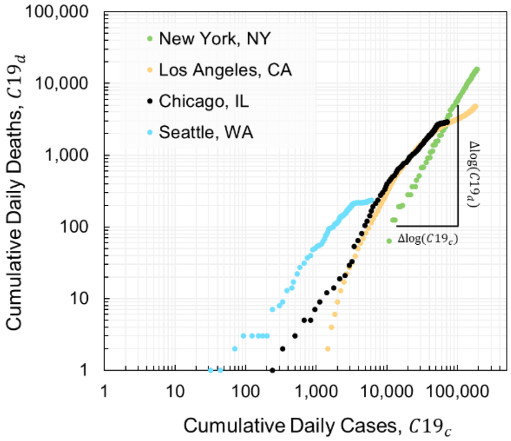


**Fig. S2**. $CFR$ distribution captured with the ratio of cumulative daily cases and deaths reported within the first 6-8 months of the COVID-19 pandemic during $\lambda_1$ for 5 US cities that were used to evaluate $CFR$ for city districts (Table S4) demonstrating the exponent in Eq.1 is close to unity.

*Urban Factors.*

The urban factors in this study are categorized into three groups: personal, indoor and outdoor. There are three main sources of input data, which come from weather, census and online maps with building footprints. From the census data (Table S1), we obtain population size, $P$, population of age 65 or greater, $N_a$, population with income of \$50,000 or less, $N_i$, and average household size $\rho_r$, which when combined with the number of buildings allows us to approximate number of housing units $(B_u)$ per building, $\rho_b$ are sufficient to derive Personal, $P_i$ and Indoor, $I_i$ Factors. To derive Outdoor Factor, $O_i$ we resort to using annual relative humidity, $H_r$ data (Table S2-S3) for the corresponding to the first wave of COVID-19 period, and building footprints, which are needed to capture 2-D planar building density and quantify city texture with an angular order between local buildings by using a 2-D order parameter, $\varphi$ (7). In the US, building footprints can oftentimes be obtained from city or state's building and planning departments (Table S2), but for locations outside the US, we resort to using OpenStreetMap (OSM) repository, since it offers a direct access to building footprints for any mapping layer visible on OSM. To extract city-wide data from OSM that would correspond to COVID-19 data, we leverage OSM boundaries depository (Table S2). However, sometimes OSM data is not complete leaving empty spaces in areas where one would expect to see multiple parcels of buildings. Thus, when using OSM data, it is important to investigate building footprints data to access its validity. To further clean and prepare data for analysis, we perform building area correction to the number of buildings. Since we are interested in measuring impact of COVID-19 in residential and community areas, we impose a minimum area of 20m² (indicative of non-residential buildings, i.e. garages) for the number of buildings, $B$ used to define corrected number of buildings, $B_a$ used in this study. Although, we were able to obtain COVID-19 data for more than 30 cities worldwide, due to building footprints limitations in countries like, India, Argentina, or Columbia, we have not been able to use them. Moreover, some OSM data provides merged buildings instead of individual ones, which underestimates values of $B_a$ and overestimates of $\rho_b$ used in determining indoor factors for Rio De Janeiro and Recife cities in Brazil, where at the city scale, one would expect to see lower values for $\rho_b$. To offset the error, we adopt average $\rho_b$ values from São Paulo, Brazil, which upon visual mapping verification, we would expect to have, on average, similar range to other major cities in Brazil.

The striking resemblance in texture between urban environments and molecular structure of polycrystalline material at an atomic scale can be established with the help of appropriate visualization techniques. To extract statistical characteristics of short and long-range city texture, we employ radial distribution function, also known as pair (or two-points) correlation function. Denoted by $g(r)$, it provides an isotropic homogenous picture of an anisotropic inhomogeneous medium by averaging the local density over time and space domains. In the context of cities and buildings, it can be thought of as a mechanism of describing density variation at a given distance from the reference building. As soon as local density deviates from the average density of a system, peaks in the distribution eventuate; in statistical particle physics terms applied to cities, this is explained as the probability of finding a building at distance $r$ from the reference building relative to randomly distributed system of buildings that at long distance converges to unity, i.e being normalized by the averaged total density of buildings within the circle defined by maximum radius, $r_{max}$. which is the limiting radius for $g(r)$ analysis—distance at which the function convergences to unity—here defined to be $15L$ where $L$ is the average building size for a city calculated using the following equation:

$$L = exp\left(\frac{1}{2N}\sum_{i=1}^{N} log(A_i)\right)$$

Eq. S1

where $N$ is the total number of buildings in the city and $A_i$ is the area of building $i$. The buildings size distributes follow a lognormal distribution. However, for many cities outside the US, it shows a significant tail for large values of L, which is representative of the fact that in older cities there are more buildings with larger areas – indicative of terraced housing. The sum of buildings projected 2-D areas divided by the city total surface gives the city built-up proportion, $\rho$. In order to quantify more accurately local average density of buildings, we utilize an average, $\rho_{city}$, from the distribution of density values as defined by:

$$\rho_{city} = exp\left(\frac{1}{N}\sum_{i=1}^{N} log\left(\frac{C_{n+1}^{r_{max}}}{\pi r_{max}^2}\right)\right)$$

Eq. S2

where $C_{n+1}^{r_{max}}$ is the total number of buildings in circular area of radius $r_{max}$. With such defined average density, $g(r)$ captures the local deviation from it in the following form:

$$g(r) = \frac{1}{N}\sum_{i=1}^{N} \frac{n_i(r+dr) - n_i(r)}{\rho_{city} 2\pi r \times dr}$$

Eq. S3

where $n_i(r)$ denotes the number of buildings within the radial distance $r$ from building $i$, and $dr$ is distance increment, which for $g(r)$ calculations we chose to be 5% of the average building size, $L$. We find that despite similar geographical location, distribution of buildings in a given ward can exhibit different characteristics of local order than that in other wards of the same city. To further quantify local texture patterns on the path of exploring meaningful ways of characterizing city texture, we enrich our city characterization toolbox with coordination number and order parameter.

In the context of buildings, coordination number, $C_n$ defines the number of neighboring buildings that are within a specific distance, $r_{min}$, from the building of reference. Although, these can be counted individually and then averaged, $C_n$ can also be obtain directly from the $g(r)$ function, using its integral in the form:

$$C_n = 2\pi\rho_{city}\int_0^{r_{min}} g(r)dr$$

Eq. S4

The approach for selecting $C_n$ should be evaluated based on its application, which depending on desired accuracy may lead to different results. Here, the application is defining average local configuration of buildings as captured by $g(r)$ thus leading to our use of Eq. S4, where $r_{min}$ is the first local minimum in the $g(r)$ distribution following the main peak (i.e. one with the maximum $g(r)$ value). Due to variability in local city texture between zip codes or wards within a given city, integral of the first peak may not always lead to $C_n > 1$, which is required to be able to obtain angular order parameter with a minimum configuration of 3 buildings, or 2 neighbors. If such situation exists, we proceed with the next local minimum in the distribution of $g(r)$ until $C_n > 1$ has been obtained. With such defined $C_n$ we proceed with calculations of the second city texture value, order parameter. Exact $r_{min}$ values used to define the first peak are presented in Tables

S4-S7 and Fig. S5c, with $g(r)$ functions visualized for cities and their first peaks for calculations of order parameters in Fig. S3.

Order parameter, $\varphi$, which is a general Mermin 2-D order parameter, designed in condensed matter physics to quantify the deviation from symmetrical order of two-dimensional crystals as:

$$\varphi = 1/N_a \left|\sum_{k=1}^{N_a} exp\ (im\vartheta_k)\right|$$

Eq. S5

where $m$ is the number of atoms in the first shell, $N_a$, the number of independent angles $\vartheta_k$ between the atom and its neighbors. For a perfectly ordered (symmetrical) system, $\varphi = 1$ for a system with a perfect m-fold orientational ordering. Applied at the city scale, $\varphi$ characterizes the average angular distortion of buildings compared to a perfect angular local order of a city at fixed $m = C_n$ within the first shell distance determined from the integral of $g(r)$ (Eq. S4):

$$\varphi = \frac{1}{N}\sum_{j=1}^{N} \frac{1}{N_a(j)}\left|\left(\sum_{k=1}^{N_a(j)} exp\ (iC_{\mathrm{n}}\vartheta_k)\right)\right|$$

Eq. S6

where, $N$ is the number of buildings. So defined, $\varphi$ = 1 represents a city in which all buildings at short distance have the same number of neighbors exhibiting angular periodicity, $2\pi/\mathrm{C_n}$; whereas any deviation from unity in this short-range city order parameter is representative of both local angular distortions of neighboring buildings, and local variations of number of neighbors that affects the number of independent angles $N_a(j)$ for each building $j = 1, N$. At the scale of cities, $\varphi$ becomes the arithmetic mean of order parameters values for all buildings. While it has been previously established that much like molecular structures cities exhibit a distinct long-range texture, which varies from gas- and liquid/glass- to crystal- like (7), here we also find that despite similar geographical location, distribution of buildings in different parts of the city (i.e. wards or districts) can exhibit different characteristics of local order (i.e. Brooklyn and Bronx in New York City, NY).

Although, gas-like (European) cities lack the expected $g(r)$ characteristics used to identify the first minimum or simply have no minima, here we utilize the distance, $r_{peak}^{g(r)}$ at which $g(r)$ reaches its first peak – a characteristic distance between one building and its nearest neighbors. This distance correlates with $r_{min}^{g(r)}$ for cities where it is possible to identify it and can be modeled using a linear correlation (8):

$$r_{min}^{g(r)} = 1.35 \times r_{peak}^{g(r)}$$

Eq. S7

Such linear correlation can be explained by the characteristic street width, which on average limits the local buildings to the nearest 2 neighbors. With such approach, we can identify $r_{min}^{g(r)}$ for any city or zip code, subsequently allowing us to derive coordination number for the first shell of neighbors. This is an important step, because distance $r_{min}^{g(r)}$ is a critical input for quantifying $\varphi$, which is used as one of the outdoor urban factor parameters.

In the present work, we seek at identifying differences in the COVID-19 spread in a given city captured through $CFR$. We therefore derive $g(r)$ functions for every location used in this work to obtain $\varphi$s, among other variable used in the determination of Urban Factors (Tables. S4-S7). However, for the calibration of weight parameters in equations 2-4 only city district ($N = 20$) from Table S3 have been used. This allowed sufficient sample of uncorrelated data from various geographical regions to be used for validation of the model ($N = 98$ for cities, zip codes and boroughs). The model parameters have been optimized by minimizing the error between predicted and measured $CFR(\lambda_1)$ values using standard statical methods. Although, it is common to apply sum of square errors when quantifying errors in regression analyses, here we adopted a sum of absolute errors, $\varepsilon$ in the form of:

$$\varepsilon = \sum_{i=1}^{n=20} |CFR_i(\lambda_1) - a_i(\lambda_1)|$$

Eq. S8

where $i$ is the city district ID from Table S4. Since $CFR$ values are fractions, differences between any predicted and measured values lead to small fractions and thus squaring the difference would lead to even smaller values thereby introducing bias to any optimization approach that is trying to minimize the sum of errors. As such the absolute sum $\varepsilon$ provides a more stable optimization parameter. We minimize $\varepsilon$ using a non-linear generalized reduced gradient (GRD) method with constraint convergence of 0.00001, forward derivates population size of 1000. With such defined parameters, we obtain optimized urban factors and intrinsic $CFR$, $a_0(\lambda_1)$ (Fig. 3).

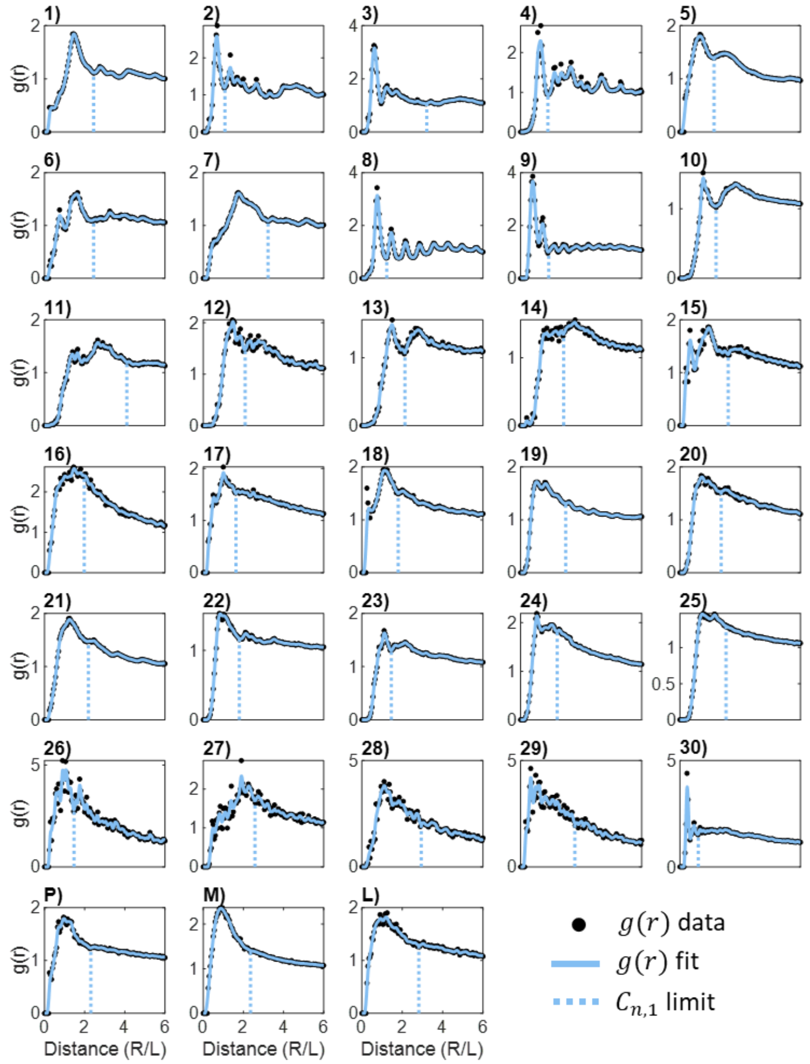


**Fig. S3**. Radial distribution function, $g(r)$, for cities in Table S5. Dashed vertical lines show the distance limits, $r_{min}$, used in the determination of order parameter, $\varphi$.

| ID | District | Zip Code | $CFR(\lambda_1)$ | $N_{a,i}/N_{0,i}$ | $P_i$ | $B$ | $B_a$ | $B_u$ | $P$ | $\rho_{r,i}$ | $\rho_{b,i}$ | $I_i$ | $R_{lim}(m)$ | $C_{n,1}$ | $\varphi$ | $\rho_i$ | $H_{R,i}$ | $O_i$ | $a_i(\lambda_1)$ |
|---|---|---|---|---|---|---|---|---|---|---|---|---|---|---|---|---|---|---|---|
| 1 | North | 60618 | 0.033 | 0.090 | 0.090 | 29,995 | 29,762 | 39,969 | 94,907 | 2.37 | 1.34 | 0.430 | 12.3 | 2.06 | 0.881 | 0.289 | 0.75 | 0.761 | 0.073 |
| 2 | North | 60625 | 0.065 | 0.100 | 0.100 | 17,939 | 17,716 | 32,451 | 79,444 | 2.45 | 1.83 | 0.528 | 14.4 | 2.34 | 0.418 | 0.266 | 0.75 | 0.727 | 0.075 |
| 3 | North | 60630 | 0.065 | 0.150 | 0.150 | 25,448 | 25,178 | 22,740 | 56,433 | 2.48 | 0.90 | 0.272 | 12.8 | 2.07 | 0.876 | 0.232 | 0.75 | 0.761 | 0.061 |
| 4 | North | 60631 | 0.100 | 0.190 | 0.190 | 16,673 | 16,271 | 12,670 | 29,529 | 2.33 | 0.78 | 0.177 | 25.9 | 6.37 | 0.401 | 0.197 | 0.75 | 0.726 | 0.047 |
| 5 | North | 60634 | 0.033 | 0.150 | 0.150 | 38,061 | 37,517 | 28,062 | 75,082 | 2.68 | 0.75 | 0.182 | 12.4 | 2.04 | 0.899 | 0.215 | 0.75 | 0.762 | 0.055 |
| 6 | North | 60641 | 0.033 | 0.120 | 0.120 | 24,540 | 24,301 | 27,131 | 69,880 | 2.58 | 1.12 | 0.378 | 12.2 | 2.03 | 0.911 | 0.294 | 0.75 | 0.763 | 0.070 |
| 7 | North | 60646 | 0.100 | 0.210 | 0.210 | 15,150 | 14,844 | 11,510 | 28,569 | 2.48 | 0.78 | 0.188 | 26.0 | 6.32 | 0.398 | 0.173 | 0.75 | 0.726 | 0.049 |
| 8 | North | 60656 | 0.030 | 0.180 | 0.180 | 11,928 | 11,568 | 12,761 | 28,218 | 2.21 | 1.10 | 0.338 | 31.9 | 9.75 | 0.362 | 0.169 | 0.75 | 0.723 | 0.057 |
| 9 | North | 60657 | 0.075 | 0.080 | 0.080 | 12,420 | 12,317 | 40,259 | 70,958 | 1.76 | 3.27 | 0.553 | 12.1 | 2.08 | 0.863 | 0.361 | 0.75 | 0.760 | 0.091 |
| 10 | Central East | 60608 | 0.030 | 0.100 | 0.100 | 17,676 | 17,392 | 28,663 | 80,059 | 2.79 | 1.65 | 0.527 | 11.9 | 2.06 | 0.821 | 0.216 | 0.65 | 0.665 | 0.063 |
| 11 | Central East | 60622 | 0.085 | 0.060 | 0.060 | 18,136 | 18,028 | 26,174 | 53,294 | 2.04 | 1.45 | 0.418 | 12.0 | 2.06 | 0.832 | 0.387 | 0.65 | 0.666 | 0.049 |
| 12 | Central East | 60623 | 0.040 | 0.110 | 0.110 | 21,975 | 21,762 | 30,324 | 81,283 | 2.68 | 1.39 | 0.468 | 12.5 | 2.07 | 0.849 | 0.234 | 0.65 | 0.667 | 0.057 |
| 13 | Central East | 60639 | 0.030 | 0.100 | 0.100 | 25,666 | 25,464 | 28,659 | 88,204 | 3.08 | 1.13 | 0.415 | 12.1 | 2.03 | 0.913 | 0.293 | 0.65 | 0.672 | 0.052 |
| 14 | Central East | 60644 | 0.065 | 0.140 | 0.140 | 11,682 | 11,589 | 19,556 | 46,591 | 2.38 | 1.69 | 0.501 | 12.7 | 2.11 | 0.885 | 0.226 | 0.65 | 0.670 | 0.064 |
| 15 | Central East | 60647 | 0.060 | 0.070 | 0.070 | 22,062 | 21,904 | 39,159 | 87,633 | 2.24 | 1.79 | 0.501 | 12.1 | 2.07 | 0.863 | 0.297 | 0.65 | 0.668 | 0.059 |
| 16 | Central East | 60651 | 0.040 | 0.120 | 0.120 | 20,088 | 20,021 | 22,726 | 63,492 | 2.79 | 1.14 | 0.401 | 12.5 | 2.04 | 0.888 | 0.293 | 0.65 | 0.670 | 0.052 |
| 17 | Central West | 60614 | 0.125 | 0.090 | 0.090 | 12,619 | 12,544 | 36,375 | 71,954 | 1.98 | 2.90 | 0.569 | 11.6 | 2.15 | 0.818 | 0.310 | 0.75 | 0.756 | 0.093 |
| 18 | Central West | 60616 | 0.070 | 0.160 | 0.160 | 8,492 | 8,398 | 26,362 | 54,197 | 2.06 | 3.14 | 0.591 | 10.9 | 2.09 | 0.873 | 0.214 | 0.75 | 0.760 | 0.109 |
| 19 | Central South | 60609 | 0.035 | 0.120 | 0.120 | 19,968 | 19,626 | 24,720 | 60,939 | 2.47 | 1.26 | 0.415 | 11.8 | 2.04 | 0.849 | 0.198 | 0.65 | 0.667 | 0.053 |
| 20 | Central South | 60621 | 0.090 | 0.140 | 0.140 | 10,747 | 10,630 | 16,072 | 28,018 | 1.74 | 1.51 | 0.386 | 13.1 | 2.05 | 0.861 | 0.157 | 0.65 | 0.668 | 0.051 |
| 21 | Central South | 60629 | 0.025 | 0.110 | 0.110 | 44,010 | 43,623 | 35,538 | 110,029 | 3.10 | 0.81 | 0.260 | 12.5 | 2.04 | 0.901 | 0.254 | 0.65 | 0.671 | 0.042 |
| 22 | Central South | 60632 | 0.027 | 0.100 | 0.100 | 30,219 | 29,868 | 27,600 | 89,857 | 3.26 | 0.92 | 0.336 | 12.5 | 2.05 | 0.881 | 0.248 | 0.65 | 0.669 | 0.046 |
| 23 | Central South | 60636 | 0.065 | 0.210 | 0.210 | 16,317 | 16,176 | 14,538 | 30,024 | 2.07 | 0.90 | 0.226 | 12.4 | 2.01 | 0.912 | 0.198 | 0.65 | 0.672 | 0.045 |
| 24 | Central South | 60638 | 0.028 | 0.140 | 0.140 | 23,540 | 32,102 | 21,239 | 58,669 | 2.76 | 0.66 | 0.108 | 13.3 | 2.10 | 0.882 | 0.225 | 0.65 | 0.670 | 0.036 |
| 25 | Central South | 60652 | 0.040 | 0.120 | 0.120 | 22,857 | 22,281 | 14,148 | 43,447 | 3.07 | 0.63 | 0.098 | 16.3 | 2.74 | 0.366 | 0.205 | 0.65 | 0.632 | 0.030 |
| 26 | South | 60617 | 0.047 | 0.150 | 0.150 | 41,314 | 40,570 | 34,492 | 83,553 | 2.42 | 0.85 | 0.235 | 13.4 | 2.11 | 0.832 | 0.113 | 0.75 | 0.757 | 0.057 |
| 27 | South | 60619 | 0.065 | 0.170 | 0.170 | 29,130 | 28,814 | 32,540 | 61,207 | 1.88 | 1.13 | 0.305 | 13.4 | 2.11 | 0.851 | 0.228 | 0.75 | 0.759 | 0.064 |
| 28 | South | 60620 | 0.065 | 0.180 | 0.180 | 32,741 | 32,372 | 30,381 | 67,711 | 2.23 | 0.94 | 0.267 | 12.9 | 2.07 | 0.882 | 0.205 | 0.75 | 0.761 | 0.063 |
| 29 | South | 60628 | 0.085 | 0.170 | 0.170 | 37,817 | 37,467 | 28,582 | 64,254 | 2.25 | 0.76 | 0.156 | 13.4 | 2.11 | 0.872 | 0.148 | 0.75 | 0.76 | 0.054 |
| 30 | South | 60643 | 0.080 | 0.190 | 0.190 | 25,311 | 24,907 | 19,799 | 48,887 | 2.47 | 0.79 | 0.202 | 26.4 | 5.68 | 0.393 | 0.179 | 0.75 | 0.725 | 0.049 |

**Table. S6.** Chicago IL, USA measured and predicted $CFR$ data with Urban Factors and input values used to derive them at the zip code level (Fig. S4a), which were merged to form Chicago districts (Fig. S1d)

| ID | District | $CFR(\lambda_1)$ | $N_{a,i}/N_{0,i}$ | $P_i$ | $B$ | $B_a$ | $B_u$ | $P$ | $\rho_{r,i}$ | $\rho_{b,i}$ | $I_i$ | $R_{lim}(m)$ | $C_{n,1}$ | $\varphi$ | $\rho_i$ | $H_{R,i}$ | $O_i$ | $a_i(\lambda_1)$ |
|---|---|---|---|---|---|---|---|---|---|---|---|---|---|---|---|---|---|---|
| 31 | Barking & Dagenham | 0.120 | 0.101 | 0.101 | 14,233 | 14,233 | 73,400 | 222,850 | 3.04 | 5.16 | 0.733 | 44.0 | 4.34 | 0.497 | 0.144 | 0.77 | 0.750 | 0.144 |
| 32 | Barnet | 0.146 | 0.135 | 0.135 | 37,683 | 37,679 | 144,020 | 414,800 | 2.88 | 3.82 | 0.692 | 24.2 | 2.35 | 0.729 | 0.127 | 0.77 | 0.767 | 0.142 |
| 33 | Bexley | 0.109 | 0.166 | 0.166 | 31,792 | 31,787 | 96,470 | 242,875 | 2.52 | 3.03 | 0.634 | 25.7 | 2.54 | 0.658 | 0.131 | 0.77 | 0.762 | 0.121 |
| 34 | Brent | 0.145 | 0.109 | 0.109 | 22,862 | 22,862 | 114,920 | 346,700 | 3.02 | 5.03 | 0.730 | 25.0 | 2.55 | 0.630 | 0.207 | 0.77 | 0.760 | 0.151 |
| 35 | Bromley | 0.113 | 0.175 | 0.175 | 52,327 | 52,322 | 137,640 | 328,225 | 2.38 | 2.63 | 0.599 | 26.1 | 2.31 | 0.343 | 0.071 | 0.77 | 0.739 | 0.099 |
| 36 | Camden | 0.120 | 0.111 | 0.111 | 7,407 | 7,407 | 106,280 | 240,425 | 2.26 | 14.35 | 0.730 | 52.9 | 5.00 | 0.423 | 0.249 | 0.77 | 0.745 | 0.140 |
| 37 | Croydon | 0.135 | 0.127 | 0.127 | 41,866 | 41,865 | 150,820 | 399,125 | 2.65 | 3.60 | 0.668 | 26.8 | 2.29 | 0.841 | 0.118 | 0.77 | 0.775 | 0.137 |
| 38 | Ealing | 0.134 | 0.112 | 0.112 | 29,151 | 29,151 | 132,310 | 367,175 | 2.78 | 4.54 | 0.704 | 24.0 | 2.55 | 0.605 | 0.187 | 0.77 | 0.758 | 0.137 |
| 39 | Enfield | 0.167 | 0.128 | 0.128 | 27,187 | 27,187 | 123,070 | 333,650 | 2.71 | 4.53 | 0.699 | 26.0 | 2.26 | 0.787 | 0.108 | 0.77 | 0.771 | 0.148 |
| 40 | Greenwich | 0.121 | 0.105 | 0.105 | 19,484 | 19,484 | 108,820 | 312,100 | 2.87 | 5.59 | 0.730 | 52.0 | 6.10 | 0.425 | 0.140 | 0.77 | 0.745 | 0.139 |
| 41 | Hammersmith & Fulham | 0.117 | 0.093 | 0.093 | 5,278 | 5,278 | 85,180 | 203,425 | 2.39 | 16.14 | 0.746 | 56.0 | 4.78 | 0.442 | 0.269 | 0.77 | 0.746 | 0.147 |
| 42 | Haringey | 0.185 | 0.090 | 0.090 | 10,907 | 10,907 | 106,770 | 283,450 | 2.65 | 9.79 | 0.750 | 62.0 | 6.16 | 0.423 | 0.208 | 0.77 | 0.745 | 0.148 |
| 43 | Harrow | 0.158 | 0.145 | 0.145 | 28,286 | 28,284 | 88,430 | 256,475 | 2.90 | 3.13 | 0.667 | 23.3 | 2.39 | 0.687 | 0.143 | 0.77 | 0.764 | 0.131 |
| 44 | Havering | 0.161 | 0.184 | 0.184 | 35,946 | 35,946 | 102,660 | 264,975 | 2.58 | 2.86 | 0.630 | 23.7 | 2.24 | 0.827 | 0.071 | 0.77 | 0.774 | 0.131 |
| 45 | Hillingdon | 0.151 | 0.130 | 0.130 | 41,564 | 41,555 | 108,570 | 298,150 | 2.75 | 2.61 | 0.627 | 24.4 | 2.60 | 0.625 | 0.098 | 0.77 | 0.759 | 0.112 |
| 46 | Hounslow | 0.116 | 0.109 | 0.109 | 25,841 | 25,835 | 99,160 | 275,825 | 2.78 | 3.84 | 0.686 | 24.8 | 2.60 | 0.590 | 0.148 | 0.77 | 0.757 | 0.128 |
| 47 | Islington | 0.142 | 0.087 | 0.087 | 5,554 | 5,554 | 105,440 | 234,725 | 2.23 | 18.98 | 0.734 | 58.6 | 5.16 | 0.454 | 0.293 | 0.77 | 0.747 | 0.140 |
| 48 | Kensington & Chelsea | 0.116 | 0.131 | 0.131 | 3,511 | 3,511 | 88,190 | 168,175 | 1.91 | 25.12 | 0.699 | 54.6 | 4.33 | 0.494 | 0.305 | 0.77 | 0.750 | 0.132 |
| 49 | Kingston upon Thames | 0.089 | 0.131 | 0.131 | 19,515 | 19,511 | 66,030 | 175,575 | 2.66 | 3.38 | 0.661 | 25.8 | 2.30 | 0.815 | 0.126 | 0.77 | 0.773 | 0.133 |
| 50 | Lambeth | 0.109 | 0.077 | 0.077 | 12,512 | 12,512 | 138,100 | 336,700 | 2.44 | 11.04 | 0.737 | 58.5 | 6.50 | 0.416 | 0.244 | 0.77 | 0.744 | 0.138 |
| 51 | Lewisham | 0.125 | 0.094 | 0.094 | 17,579 | 17,579 | 122,300 | 310,550 | 2.54 | 6.96 | 0.721 | 52.5 | 5.89 | 0.395 | 0.188 | 0.77 | 0.743 | 0.132 |
| 52 | Merton | 0.117 | 0.120 | 0.120 | 18,020 | 18,020 | 82,900 | 212,075 | 2.56 | 4.60 | 0.689 | 45.9 | 5.36 | 0.453 | 0.163 | 0.77 | 0.747 | 0.124 |
| 53 | Newham | 0.124 | 0.067 | 0.067 | 11,756 | 11,756 | 109,650 | 370,100 | 3.38 | 9.33 | 0.791 | 43.3 | 3.77 | 0.476 | 0.193 | 0.77 | 0.749 | 0.176 |
| 54 | Redbridge | 0.144 | 0.121 | 0.121 | 25,436 | 25,430 | 102,320 | 310,000 | 3.03 | 4.02 | 0.708 | 44.7 | 5.23 | 0.460 | 0.143 | 0.77 | 0.747 | 0.133 |
| 55 | Richmond upon Thames | 0.140 | 0.143 | 0.143 | 20,579 | 20,577 | 83,180 | 195,700 | 2.35 | 4.04 | 0.657 | 25.4 | 2.29 | 0.796 | 0.097 | 0.77 | 0.772 | 0.132 |
| 56 | Southwark | 0.087 | 0.078 | 0.078 | 11,203 | 11,203 | 132,590 | 338,225 | 2.55 | 11.84 | 0.750 | 54.0 | 5.17 | 0.470 | 0.212 | 0.77 | 0.748 | 0.148 |
| 57 | Sutton | 0.089 | 0.150 | 0.150 | 23,785 | 23,778 | 81,390 | 201,175 | 2.47 | 3.42 | 0.647 | 26.4 | 2.32 | 0.792 | 0.128 | 0.77 | 0.772 | 0.130 |
| 58 | Tower Hamlets | 0.122 | 0.059 | 0.059 | 6,474 | 6,474 | 119,340 | 326,625 | 2.74 | 18.43 | 0.778 | 51.3 | 3.94 | 0.472 | 0.222 | 0.77 | 0.748 | 0.164 |
| 59 | Waltham Forest | 0.12 | 0.101 | 0.101 | 15,169 | 15,169 | 101,140 | 283,175 | 2.80 | 6.67 | 0.738 | 53.0 | 6.00 | 0.395 | 0.162 | 0.77 | 0.743 | 0.142 |
| 60 | Wandsworth | 0.1 | 0.089 | 0.089 | 12,305 | 12,305 | 138,880 | 346,225 | 2.49 | 11.29 | 0.743 | 60.8 | 6.17 | 0.416 | 0.213 | 0.77 | 0.744 | 0.143 |

**Table. S7.** London UK measured and predicted $CFR$ data with Urban Factors and input values used to derive them at the level of boroughs (Fig. S4b).

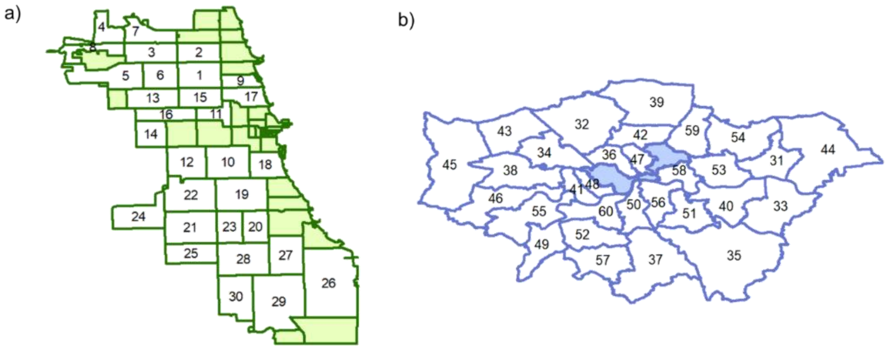


**Fig. S4**. (a) Chicago IL, USA zip codes and (b) Greater London UK boroughs.

To verify the validity of the model and its multi-spatial scale global application, we test the correlation between predicted and measured population adjusted $CFR_{pop}$ values for 118 location worldwide (Fig. 4b). Although, we find that $CFR_{pop}(\lambda_1)$ for several cities can't be predicted using the model, we identify those as places either isolated territories (island or small countries) or regions that were quick to adopt social distancing and any lockdown measures during the first wave, $\lambda_1$, of the COVID-19 pandemic. It is worth noting that any country used in this study for model verification as listed in table S5, due to its small population size and area, is treated as a city and thus compared to any other city listed in the table. To further study verify the multi scale application, we introduce data for 30 zip codes from Chicago IL, USA (Fig. S4a) and 30 boroughs from Greater London region in the UK (Fig. S4b) as well as 8 wards from Washington D.C. USA (Fig. S5), which due to their low population values ($< 100{,}000$) were not used in the optimization of the model in Eq. 2. Not only this data presents that measured $CFR_{pop}(\lambda_1)$ varies significantly across different locations, but it also shows that the Urban Factors model can predict accurately $CFR_{pop}$ values (Fig. 4b) at the scale smaller than city and district levels.

a)

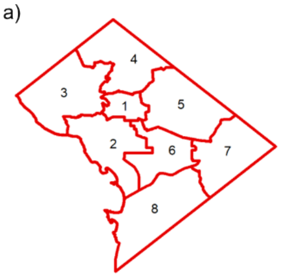


b)

| City | ID-SI | District | $CFR_s(\lambda_1)$ | $CFR(\lambda_1)$ | $CFR_m(\lambda_1)$ | $N_{a,i}/N_{0,i}$ | $N_{I,i}/N_{0,i}$ | $P_i$ | $B$ | $B_a$ | $B_u$ | $P$ | $\rho_{r,i}$ | $\rho_{b,i}$ | $I_i$ | $R_{lim}(m)$ | $C_{n,1}$ | $\varphi$ | $\rho_i$ | $H_{R,i}$ | $O_i$ | $a_i(\lambda_1)$ |
|---|---|---|---|---|---|---|---|---|---|---|---|---|---|---|---|---|---|---|---|---|---|---|
| Washington D.C. | 1 | Ward 1 | 0.047 | 0.040 | 0.042 | 0.070 | 0.25 | 0.070 | 12,462 | 12,049 | 39,068 | 83,811 | 2.15 | 3.24 | 0.607 | 9.1 | 2.14 | 0.837 | 0.298 | 0.65 | 0.659 | 0.0668 |
| | 2 | Ward 2 | 0.084 | 0.056 | 0.058 | 0.100 | 0.22 | 0.100 | 11,714 | 11,160 | 42,701 | 77,855 | 1.82 | 3.83 | 0.584 | 10.1 | 2.35 | 0.811 | 0.178 | 0.65 | 0.657 | 0.0637 |
| | 3 | Ward 3 | 0.110 | 0.068 | 0.075 | 0.180 | 0.17 | 0.180 | 21,565 | 19,888 | 40,665 | 82,737 | 2.03 | 2.04 | 0.510 | 21.0 | 3.77 | 0.607 | 0.130 | 0.65 | 0.643 | 0.0645 |
| | 4 | Ward 4 | 0.038 | 0.036 | 0.039 | 0.050 | 0.28 | 0.050 | 29,771 | 26,923 | 33,459 | 89,992 | 2.69 | 1.24 | 0.429 | 13.9 | 3.03 | 0.483 | 0.148 | 0.65 | 0.633 | 0.0484 |
| | 5 | Ward 5 | 0.062 | 0.056 | 0.057 | 0.140 | 0.38 | 0.140 | 26,836 | 24,607 | 38,521 | 90,172 | 2.34 | 1.57 | 0.475 | 13.4 | 2.99 | 0.690 | 0.152 | 0.65 | 0.649 | 0.0547 |
| | 6 | Ward 6 | 0.044 | 0.061 | 0.069 | 0.100 | 0.23 | 0.100 | 23,242 | 21,937 | 51,191 | 99,786 | 1.95 | 2.33 | 0.526 | 8.7 | 2.16 | 0.507 | 0.243 | 0.65 | 0.635 | 0.0572 |
| | 7 | Ward 7 | 0.064 | 0.056 | 0.056 | 0.130 | 0.53 | 0.130 | 22,735 | 19,720 | 34,727 | 81,946 | 2.36 | 1.76 | 0.510 | 24.0 | 5.17 | 0.454 | 0.113 | 0.65 | 0.631 | 0.0575 |
| | 8 | Ward 8 | 0.072 | 0.079 | 0.085 | 0.090 | 0.63 | 0.090 | 15,011 | 14,148 | 34,844 | 86,384 | 2.48 | 2.46 | 0.595 | 22.6 | 4.37 | 0.536 | 0.091 | 0.65 | 0.637 | 0.0660 |

**Fig. S5**. Washington D.C. USA wards data showing a) geographical boundaries of wards and b) measured and predicted $CFR$ with their input values.

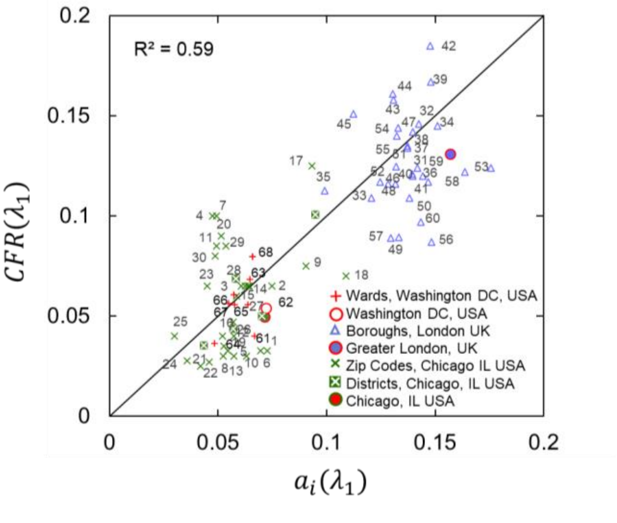


**Fig. S6**. Urban Factors model predications showing a comparison between measured and predicted $CFR(\lambda_1)$ from Eq.5 for 30 zip codes in Chicago IL, USA (Table S6) and 30 boroughs in London, UK (Table S7), 8 wards in Washington D.C. USA between 03/2020-08/2020. Linear fitting with slope coefficient of unity provides $R^2 = 0.59$ and $RMSE = 0.027$. For comparison city and city district data is presented in the figure. Predicted $CFR$ values used the urban factor weight parameters from figure 3, the same as $CFR$ values in figure 4.

**References**

1. A. L. Hill, The math behind epidemics. *Physics Today* **73**, 28–34 (2020).

2. G. Fan, *et al.*, Decreased Case Fatality Rate of COVID-19 in the Second Wave: A study in 53 countries or regions. *Transbound Emerg Dis* **68**, 213–215 (2021).

3. C.-S. Chang, *et al.*, The computation of case fatality rate for novel coronavirus (COVID-19) based on Bayes theorem. *Medicine (Baltimore)* **99**, e19925 (2020).

4. S. Gupta, K. Kumar Patel, S. Sivaraman, A. Mangal, Global Epidemiology of First 90 Days into COVID-19 Pandemic: Disease Incidence, Prevalence, Case Fatality Rate and Their Association with Population Density, Urbanisation and Elderly Population. *Journal of Health Management* **22**, 117–128 (2020).

5. C. S. Narayanan, A novel cohort analysis approach to determining the case fatality rate of COVID-19 and other infectious diseases. *PLOS ONE* **15**, e0233146 (2020).

6. M. Neil, N. Fenton, M. Osman, S. McLachlan, Bayesian network analysis of Covid-19 data reveals higher infection prevalence rates and lower fatality rates than widely reported. *Journal of Risk Research* **23**, 866–879 (2020).

7. J. M. Sobstyl, T. Emig, M. J. A. Qomi, F.-J. Ulm, R. J.-M. Pellenq, Role of City Texture in Urban Heat Islands at Nighttime. *Phys. Rev. Lett.* **120**, 108701 (2018).

8. J. Roxon, “Role of city texture in identifying drag coefficients of buildings to prevent hurricane damage,” Massachusetts Institute of Technology. (2020).
